# Validating and modeling the impact of high-frequency rapid antigen screening on COVID-19 spread and outcomes

**DOI:** 10.1101/2020.09.01.20184713

**Authors:** Beatrice Nash, Anthony Badea, Ankita Reddy, Miguel Bosch, Nol Salcedo, Adam R. Gomez, Alice Versiani, Gislaine Celestino Dutra Silva, Thayza Maria Izabel Lopes dos Santos, Bruno H. G. A. Milhim, Marilia M Moraes, Guilherme Rodrigues Fernandes Campos, Flávia Quieroz, Andreia Francesli Negri Reis, Mauricio L. Nogueira, Elena N. Naumova, Irene Bosch, Bobby Brooke Herrera

## Abstract

High frequency screening of populations has been proposed as a strategy in facilitating control of the COVID-19 pandemic. We use computational modeling, coupled with clinical data from rapid antigen tests, to predict the impact of frequent viral antigen rapid testing on COVID-19 spread and outcomes. Using patient nasal or nasopharyngeal swab specimens, we demonstrate that the sensitivity/specificity of two rapid antigen tests compared to quantitative real-time polymerase chain reaction (qRT-PCR) are 82.0%/100% and 84.7%/85.7%, respectively; moreover, sensitivity correlates directly with viral load. Based on COVID-19 data from three regions in the United States and São José do Rio Preto, Brazil, we show that high frequency, strategic population-wide rapid testing, even at varied accuracy levels, diminishes COVID-19 infections, hospitalizations, and deaths at a fraction of the cost of nucleic acid detection via qRT-PCR. We propose large-scale antigen-based surveillance as a viable strategy to control SARS-CoV-2 spread and to enable societal re-opening.

## INTRODUCTION

The COVID-19 pandemic has taken an unprecedented toll on lives, wellbeing, healthcare systems, and global economies. As of 13 April 2021, there have been more than 136.2 million confirmed cases globally with over 2.9 million confirmed deaths ^1^. However, these statistics and the current mapping of disease spread present an incomplete picture of the outbreak largely due to the lack of adequate testing, particularly as undetected infected cases are the main source of disease spread ^2–7^. It is estimated that the number of infected cases is more than 6 times greater than the cases reported^8^. As of April 2021, the United States, India, and Brazil remain the top three countries with the highest number of COVID-19 cases and deaths worldwide. As countries begin to re-open their economies, a method for accessible and frequent surveillance of COVID-19, with the necessary rapid quarantine measures, is crucial to prevent the multiple resurgences of the disease.

The current standard of care rightfully places a strong focus on the diagnostic limit of detection, yet frequently at the expense of cost and turnaround time. This approach has contributed to limited population testing largely due to a dearth of diagnostic resources. Quantitative real-time polymerase chain reaction (qRT-PCR) is the gold-standard method for clinical diagnosis, with high sensitivity and specificity, but these tests require trained personnel, expensive reagents and instrumentation, and significant time to execute^9, 10^. Facilities offering qRT-PCR sometimes require a week or longer to complete and return the results to the patient^11, 12^. During this waiting period the undiagnosed individual may spread the infection and/or receive delayed medical treatment. Moreover, due to the cost and relative inaccessibility of qRT-PCR in both resource-limited and abundant settings, large-scale screening using qRT-PCR at frequent intervals remains impractical to identify infected but asymptomatic or mildly symptomatic infections. Numerous studies have reported asymptomatic SARS-CoV-2 infections as well as a variation in viral load within and between individuals at different time points, suggesting the need for more frequent testing for informative surveillance^13–18^.

Technologies such as rapid viral antigen detection, clustered regularly interspaced short palindromic repeats (CRISPR), and loop-mediated isothermal amplification (LAMP) of SARS-CoV-2 provide potential large-scale screening applications, yet their implementation is stymied by requirements for qRT-PCR-like accuracy before they can reach the market ^19^. Several members of the scientific and medical community have emphasized the value of widespread and frequent antigen testing^20–22^. In countries such as India, where the qRT-PCR resources would not be sufficient to cover monitoring of the population, the use of rapid antigen tests is well underway^23, 24^. In early May 2020, the United States Food and Drug Administration (FDA) authorized the first antigen test for the laboratory detection of COVID-19, citing a need for testing beyond molecular and serological methods. Antigen testing detects the viral proteins rather than nucleic acids or human antibodies, allowing for detection of an active infection with relative ease of sample collection and assay. These rapid assays – like other commercially-available rapid antigen tests - can be mass-produced at low prices and be administered by the average person without a laboratory or instrumentation. These tests also take as little as 15 minutes to determine the result, enabling real-time diagnosis and/or surveillance. Although antigen tests usually perform with high specificities (true negative rate), their sensitivity (true positive rate) is often lower when compared to molecular assays. While qRT-PCR can reach a limit of detection as low as 10^2^ genome copies per mL, rapid antigen testing detects viral protein that is assumed to correlate with approximately 10^5^ genome copies per mL ^25^.

We hypothesize that frequent antigen-based rapid testing even with lower sensitivities compared to qRT-PCR - along with appropriate quarantine measures - can be more effective at decreasing COVID-19 spread than less frequent molecular testing of symptomatic individuals. Keeping in mind the realities of daily testing in resource-limited regions, we also hypothesize that testing frequency can be adjusted according to the prevalence of the disease; that is, an uptick in reported cases should be accompanied by more frequent testing. During the viral incubation period, high infectivity correlates with a high viral load that can be detected by either qRT-PCR or rapid antigen testing ^18, 21, 26–28^. Rapid tests thus optimize diagnosis for the most infectious individuals. Studies also point to the relatively small window of time during an individual’s incubation period in which the qRT-PCR assay is more sensitive than rapid tests ^21^.

In this study we report the clinical validation of two direct antigen rapid tests for detection of SARS-CoV-2 spike glycoprotein (S) or nucleocapsid protein (N) using retrospectively collected nasopharyngeal or nasal swab specimens. Using the clinical performance data, we develop a modeling system to evaluate the impact of frequent rapid testing on COVID-19 spread and outcomes using a variation of a SIR model, which has been previously used to model COVID-19 transmission ^29–35^. We build on this model to incorporate quarantine states and testing protocols to examine the effects of different testing regimes. This model distinguishes between undetected and detected infections and separates severe cases, specifically those requiring hospitalization from those less so, which is important for disease response systems such as intensive care unit triaging. We simulate COVID-19 spread with rapid testing and model disease outcomes in three regions in the United States and São José do Rio Preto, Brazil - the site of the clinical validation study - using publicly available data. To date, COVID-19 modeling describes the course of disease spread in response to social distancing and quarantine measures, and a previous simulation study has shown that frequent testing with accuracies less than qRT-PCR, coupled with quarantine process and social distancing, are predicted to significantly decrease infections ^21, 29, 35–39^. Godio et al. and Hou et al. use the classic SEIR model to predict COVID-19 spread and dynamics. Reno et al. use this approach to predict COVID-19-associated hospitalizations under social distancing policies. While SEIR models are foundational for epidemiological studies, they fail to distinguish between diagnosed and undiagnosed individuals. To address this limitation, Giordino et al. propose and implement a SIDHARTHE model, on which our model is based, to understand disease spread in Italy, but their analysis does not incorporate different testing regimes. Larremore et al. discuss how rapid testing strategies, even when applied with low-sensitivity tests, are useful, as we do, when applied to their S-I-R-Q-SQ model. Neither Giordino et al. nor Larremore et al. use realized outbreaks to extract parameters - instead they are chosen based on informed guesses and/or idealized closed systems. They also do not compare the results of their simulations with the data reported, and hence the analyses are limited by their purely theoretical nature.

By simulating the implementation of rapid testing strategies using parameters extracted from data from realized outbreaks, we are able to expand on existing insights, including: predicting the effectiveness of such schema on outbreaks with differing dynamics and at varying intervention times, extracting parameters to train the comprehensive SIDHARTHE-Q model, and demonstrating a method that is easily applied to fit parameters for any COVID-19 outbreak given a data set including daily reports of confirmed cases, current hospitalizations, and deaths. Using this method, we propose and test the effectiveness of a variety of testing strategies and analyze key factors affecting their success or failure. Both simulations and data-driven predictions are of utmost importance to make decisions concerning an unprecedented event such as a rapidly-evolving pandemic, and this is the first modeling system using publicly-available data to simulate how potential public health strategies based on testing performance, frequency, and geography impact the course of COVID-19 spread and outcomes.

Our findings suggest that a rapid test, even with sensitivities lower than molecular tests, when strategically administered 2-3 times per week, will reduce COVID-19 spread, hospitalizations, and deaths at a fraction of the cost of nucleic acid testing via qRT-PCR. Modern surveillance systems should be well equipped with rapid testing tools to ensure that disease tracking and control protocols are effective and well-tailored to national, regional, and community needs.

## RESULTS

### Accuracy of Direct Antigen Rapid Tests Correlate with Viral Load Levels

Rapid antigen tests have recently been considered a viable source for first-line screening, although concerns regarding the accuracy of these tests persist. We clinically validated two different direct antigen rapid tests for the detection of either nucleocapsid protein (N) or spike glycoprotein (S) from SARS-CoV-2 in retrospectively collected nasal or nasopharyngeal swab specimens. Of the total number of nasal swab specimens evaluated by qRT-PCR for amplification of SARS-CoV-2 N, S, and ORF1ab genes, 100 tested positive and 58 tested negative (Table 1). The overall sensitivity and specificity of the rapid antigen test for detection of SARS-CoV-2 N, evaluated across the nasal swab specimens, was 82.0% and 100%, respectively. Of the total number of nasopharyngeal swab specimens evaluated by qRT-PCR for amplification of SARS-CoV-2 N, RNA-dependent RNA polymerase (RdRp), and envelope (E) genes, 72 tested positive and 49 tested negative (Table 2). The overall sensitivity and specificity of the rapid antigen test for detection of SARS-CoV-2 S, evaluated across the nasopharyngeal swab specimens was 84.7% and 85.7%, respectively.

**Table 1.** Clinical validation summary for the direct antigen rapid test (DART) for SARS-CoV-2 nucleocapsid protein evaluated using 158 retrospectively collected patient nasal swab specimens.

| All Data Summary |  |  |  |  |  |  |  |  |
| --- | --- | --- | --- | --- | --- | --- | --- | --- |
|  |  | qRT-PCR<br>(gene average) |  |  |  |  | 95% Confidence<br>Interval |  |
|  |  | + | - | Total | Sensitivity | 82.0% | 73.1% | 88.9% |
| DART | + | 82 | 0 | 82 | Specificity | 100.0% | 93.8% | 100.0% |
|  | - | 18 | 58 | 76 | Positive<br>Predictive<br>Value | 100.0% |  |  |
| Total |  | 100 | 58 | 158 | Negative<br>Predictive<br>Value | 76.3% | 67.9% | 83.0% |
|  |  |  |  |  | Prevalence | 63.3% | 55.3% | 70.8% |
|  |  |  |  |  | Overall<br>Agreement | 88.6% | 82.6% | 93.1% |

**Table 2.** Clinical validation summary for the SARS-CoV-2 direct antigen rapid test (DART) for SARS-SoC-2 spike glycoprotein evaluated using 121 retrospectively collected patient nasopharyngeal swab specimens.

| All Data Summary |  |  |  |  |  |  |  |  |
| --- | --- | --- | --- | --- | --- | --- | --- | --- |
|  |  | qRT-PCR<br>(gene average) |  |  |  |  | 95% Confidence<br>Interval |  |
|  |  | + | - | Total | Sensitivity | 84.7% | 74.3% | 92.1% |
| DART | + | 61 | 7 | 68 | Specificity | 85.7% | 72.8% | 94.1% |
|  | - | 11 | 42 | 53 | Positive<br>Predictive<br>Value | 89.7% | 81.3% | 94.6% |
| Total |  | 72 | 49 | 121 | Negative<br>Predictive<br>Value | 79.3% | 68.7% | 86.9% |
|  |  |  |  |  | Prevalence | 59.5% | 50.2% | 68.3% |
|  |  |  |  |  | Overall<br>Agreement | 85.1% | 77.5% | 90.9% |

The Ct value indirectly quantifies the viral RNA copy number related to the viral load of the sample for the specific assay ^40–42^. Ct values represent the number of qRT-PCR cycles at which generated fluorescence crosses a threshold during the linear amplification phase; Ct values are therefore inversely related to the viral load. Our data demonstrate that the sensitivity of the rapid antigen tests are positively correlated to the viral load level (Table 3). For the SARS-CoV-2 N and S rapid tests, the sensitivities were greater than 90% when tested with samples containing Ct values <25, but plateaued to approximately 80-85% when tested with samples containing Ct values between 30-40 (Table 3, Supplementary Fig.1). Taken together, the clinical data shows that the rapid antigen test performs with increasing accuracy for individuals with a higher viral load, and potentially the most infectious ^18, 26–28^.

**Fig. 1.**
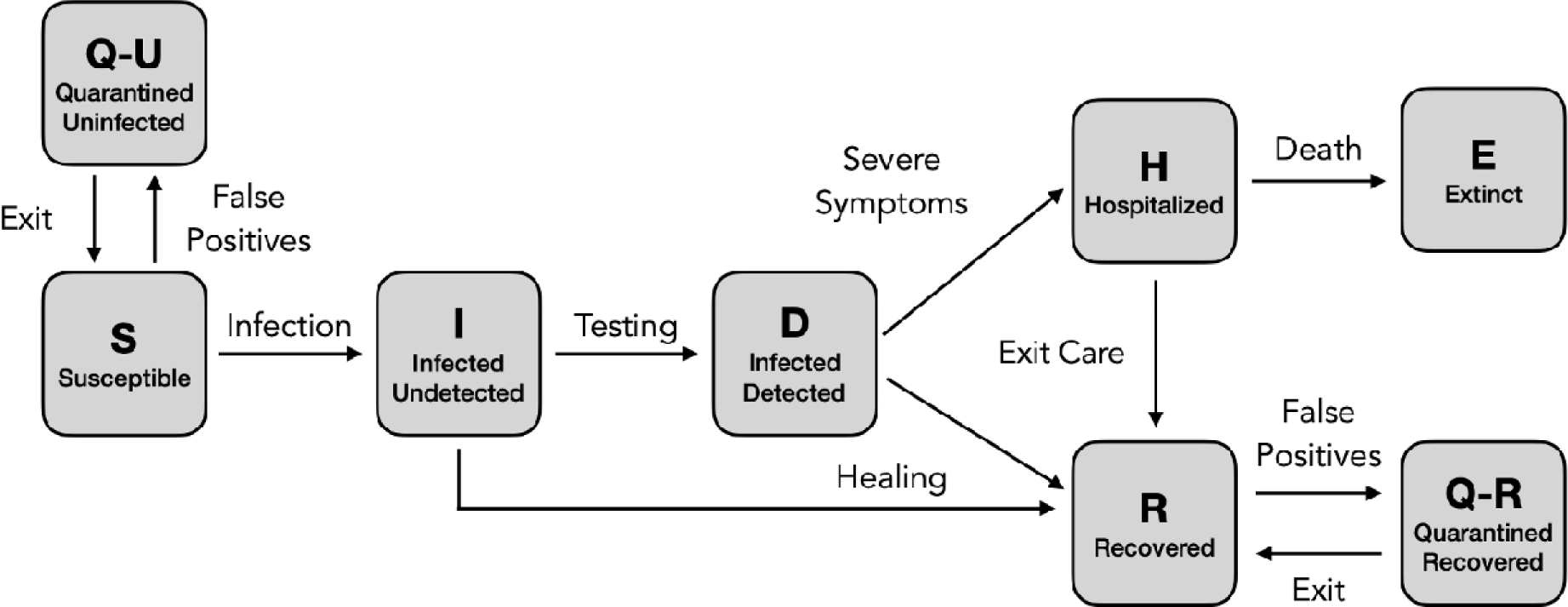
Graphical scheme displaying the relationships between the stages of quarantine and infection in *SIDHRE-Q* model. **Q-U**, quarantine uninfected; **S**, susceptible (uninfected); **I**, infected undetected (pre-testing and infected); **D**, infected detected (infection diagnosis through testing); **H**, hospitalized (infected with life threatening symptom progression); **R**, recovered (healed); **E**, extinct (dead); and **Q-R**, quarantine recovered (healed but in quarantine by false positive testing).

**Table 3.** Data summary of direct antigen rapid test (DART) for detection of SARS-CoV-2 nucleocapsid protein and DART for detection of SARS-CoV-2 spike glycoprotein performance in comparison to qRT-PCR results. Sensitivity, specificity, Positive predicative value, (PPV) negative predictive value (NPV), prevalence, and overall agreement are calculated for increasing PCR cycle threshold (Ct) values.

|  |  | Cycle threshold (Ct) value | Total Cases | DART Positives | DART Negatives | Sensitivity | Specificity |
| --- | --- | --- | --- | --- | --- | --- | --- |
| DART (nucleocapsid protein) | SARS-CoV-2 (Gene Average) | < 20 | 82 | 24 | 58 | 95.8% | 100.0% |
|  |  | < 25 | 111 | 53 | 58 | 90.6% | 100.0% |
|  |  | < 30 | 136 | 78 | 58 | 83.3% | 100.0% |
|  |  | < 35 | 157 | 99 | 58 | 79.8% | 100.0% |
|  |  | < 40 | 158 | 100 | 58 | 80.0% | 100.0% |
| DART (spike glycoprotein) | SARS-CoV-2 (Gene Average) | < 10 | 51 | 2 | 49 | 100.0% | 85.7% |
|  |  | < 15 | 72 | 23 | 49 | 100.0% | 85.7% |
|  |  | < 20 | 90 | 41 | 49 | 97.6% | 85.7% |
|  |  | < 25 | 111 | 62 | 49 | 91.9% | 85.7% |
|  |  | < 30 | 121 | 72 | 49 | 84.7% | 85.7% |
|  |  | < 35 | 121 | 72 | 49 | 84.7% | 85.7% |

### An Enhanced Epidemiological SIDHRE-Q Model

We propose an enhanced epidemiological modeling system, *SIDHRE-Q,* a variant of the classical SIR model in order to expand our clinical validation study and to understand the effects of using frequent rapid tests such as the rapid antigen test on COVID-19 outbreak dynamics. The changes we make to the basic model to encompass the unique characteristics of the COVID-19 pandemic are similar to those presented by Giordano et al.^29^ (Fig. 1, Supplementary Fig. 2). The differential equations governing the evolution of the *SIDHRE-Q* model and descriptions of the parameter values are provided in the methods section (Equation 1, Table 4).

**Table 4.** Details of parameter values used for *SIDHRE-Q* Model.

| Parameter | Details & Statistics |  |  |  |
| --- | --- | --- | --- | --- |
| $\alpha$ | $\alpha$ is rate of transmission between **I** and **S**. It is defined as, per day, the probability that an interaction between an undetected infected person and an uninfected person results in a new infection, multiplied by the average number of uninfected people an undetected infected person comes into contact in a day. $\alpha$ is estimated from the data. Units: 1/(days). | | Mean | St. Dev. |
|  |  | MA | 0.088 | 0.051 |
|  |  | LA | 0.090 | 0.034 |
|  |  | NYC | 0.067 | 0.042 |
|  |  | SJRP | 0.121 | 0.042 |
| $\eta$ | $\eta$ is rate of transmission between **D** and **S**. It is defined as the probability that an interaction between an infected person and an uninfected person results in a new infection, multiplied by the average number of uninfected people a detected infected person comes into contact with in a given day. Units: 1/(days).<br>$\eta = 0.01 \cdot \alpha$<br>The constant relating $\eta, \alpha$ accounts for a small but nonzero transmission due to the quarantined (detected) infected population. This value was chosen to be small, assuming an individual in a mandated quarantine will only interact with others with low probability, such as within a household, where complete isolation is difficult to maintain. | | | |
| $\nu$ | $\nu$ is symptomatic detection rate. It is defined as the probability that a symptomatic undetected individual is diagnosed per day. $\nu$ is estimated from the data. $\nu$ is multiplied by sensitivity (assume benchmark sensitivity 100% for PCR, as used when fitting). Units: 1/days. | | Mean | St. Dev. |
|  |  | MA | 0.006 | 0.005 |
|  |  | LA | 0.011 | 0.006 |
|  |  | NYC | 0.0056 | 0.002 |
|  |  | SJRP | 0.015 | 0.007 |
| $\epsilon$ | $\epsilon$ is asymptomatic detection rate. It is defined as the probability that an asymptomatic undetected infected individual is diagnosed on a given day. $\epsilon = 0$ while fitting (during PCR symptomatic testing). $\epsilon = (\text{sensitivity}/\text{days between tests})$ when the rapid testing strategy is activated. Units: 1/days. | | | |
| $\lambda$ | $\lambda$ is undetected recovery rate. It is defined as the probability that an undetected infected individual transitions to the recovered state on a given day. $\lambda = 1/10$, or the inverse of average recovery time<sup>72</sup>. Units: 1/days. | | | |
| $\mu$ | $\mu$ is rate of onset of severe symptoms. It is the probability that an infected individual develops severe symptoms on a given day and transitions into the hospitalized state. The flow from **D** to **H** is assumed to be independent of the ratio $I/D$, but comes only from the detected infected population, hence why it is multiplied by $(I + D)/D$. $\mu$ is estimated from the data. Units: 1/days. | | Mean | St. Dev. |
|  |  | MA | 0.0013 | 9.5e-4 |
|  |  | LA | 0.0016 | 2.4e-4 |
|  |  | NYC | 0.0011 | 6.6e-4 |
|  |  | SJRP | 0.0018 | 8.0e-4 |
| $\rho$ | $\rho$ is detected recovery rate. It is the probability that a detected infected individual transitions to the recovered state on a given day.<br>$\rho = 1/10$, or the inverse of the average recovery time<sup>72</sup>. Units: 1/days. | | | |
| $\sigma$ | $\sigma$ is the hospitalized recovery rate. It is the probability that a hospitalized individual transitions to the recovered state on a given day. $\sigma = 1/11$, or the inverse of the average recovery time for a hospitalized individual<sup>72</sup>. Units: 1/days. | | | |
| $\tau$ | $\tau$ is hospitalized rate of death. It is the probability that a hospitalized individual expires on a given day. $\tau$ is estimated from the data. Units: 1/days. | | Mean | St. Dev. |
|  |  | MA | 0.034 | 0.012 |
|  |  | LA | 0.016 | 0.004 |
|  |  | NYC | 0.036 | 0.034 |
|  |  | SJRP | 0.032 | 0.045 |
| $\gamma$ | $\gamma$ is the false positive rate. It is the probability of entering either of the quarantine states on a given day from either the Susceptible or Recovered populations. $\gamma = 0$ while fitting (during PCR symptomatic testing). $\gamma = (1 - \text{specificity}) \times (1/\text{days between tests})$ when the rapid testing strategy is activated. Units: 1/days. | | | |
| $\psi$ | $\psi$ is the rate of exit from quarantine. It is the probability that an individual exits quarantine on a given day. $\psi = 1/10$, or the inverse of the quarantine period for fixed length quarantine. Units: 1/days. | | | |

An individual that begins in Susceptible (**S)** may either transition to a Quarantine Uninfected (**Q-U**) state via a false positive result or to an Infected Undetected (**I**) state via interaction with an infected individual. Should an individual in **S** move into **Q-U**, they are quarantined for 10 days before returning to **S**, a time period chosen based on current knowledge of the infectious period of the disease and is consistent with CDC guidelines^43^. One could also conceive of an effective strategy in which individuals exit quarantine after producing a certain number of negative rapid tests in the days following their initial positive result or confirm their negative result using qRT-PCR. Prolonged incubation beyond 10 days is assumed to be unlikely – post-quarantine risk of transmission is estimated at 1% - and hence is not included in this probabilistic model. ^43^

State **I** contains individuals who are infected but not diagnosed. Given that those diagnosed are predominantly quarantined, the undiagnosed individuals in **I** – many of which are pre- or asymptomatic – interact more with the **S** population than do those in Infected Detected (**D**) and transmission due to this population is critically important to modeling outbreaks. Therefore, the infectious rate for **I** is assumed to be significantly larger than for **D**. Furthermore, a region’s ability to control an outbreak is directly related to how quickly and effectively the population in **I** tests into **D**, reducing transmission rates through quarantine. From both **I** and **D** individuals may transition into Recovered (**R)**, accounting for the many cases of infection that are never detected. This study, in particular, highlights the critical role frequency of testing, along with strict quarantine, has in mitigating the spread of the disease and provides specific testing strategies based on rapid tests we predict to be highly effective.

In this model, we assume that individuals receive a positive diagnosis before developing severe symptoms and that those with symptoms severe enough to be potentially fatal will go to the hospital. If an individual develops symptoms, we assume they are tested daily until receiving a positive result; hence, before severe symptoms develop, they will be diagnosed with high probability. Those who do not develop symptoms are tested according to the frequency of tests administered to the general population. Therefore, there is no modeled connection between **I** and Hospitalized (**H)** or between **I** and Extinct (**E**), i.e. dead. Removing these assumptions would have negligible impact on the results as these flows are very small. We estimated the flows using data on approximate total deaths due to COVID based on excess deaths in the states examined and found them to be zero for greater than 10% of the days considered for each location. Although lacking this information for São José do Rio Preto, we made the same assumption.

Should an individual test positive and transition to **D**, they may either develop serious symptoms requiring care or recover. Those who develop serious symptoms and transition to state **H** will then transition to either **R** or **E**. The recovered population is also tested with the same frequency as the rest of the population, as infected individuals may recover without being detected and the modeled testing strategy has no way of differentiating with certainty between false positives and true positive, asymptomatic cases. Therefore, the Quarantined Recovered (**Q-R**) state is introduced with the same connections to **R** as the connections between **S** and **Q-U**. Though the reinfection rate of SARS-CoV-2 has been a point of recent debate, it is assumed that the number of re-infected individuals is small ^44–48^. Therefore, individuals cannot transition from **R** to **S**, hence the separately categorized quarantined populations. As further knowledge regarding reinfection rate develops as the pandemic continues, a flow could be added from **R** to **S** with rate inversely proportional to the time for which immunity lasts.

We considered several variations and extensions of the *SIDHRE-Q* model. In simulations, we tested additional states, such as those in the *SIDARTHE* model, which include distinctions between symptomatic and asymptomatic cases for both detected and undetected populations^29^. Correlations between viral load and infectivity and sensitivity were also considered. Altogether, our modeling system has been well tuned to predict the impact of high frequency rapid testing on current COVID-19 spread and outcomes.

### Frequent Rapid Testing with Actionable Quarantining Dramatically Reduces Disease Spread

In order to demonstrate how strategies could affect the disease spread in different geographies and demographics, we used surveillance data obtained from regions of varying characteristics: the state of Massachusetts (MA), New York City (NYC), Los Angeles (LA), and São José do Rio Preto (SJRP), Brazil, the site of the rapid antigen test clinical validation study. These regions are also selected in our study due to the readily available surveillance data provided by the local governments. We fit the model to the data from each region starting 1 April 2020. At this time point the disease reportedly is most advanced in NYC and least advanced in SJRP, Brazil with estimated cumulative infection rates of 7.11% and 0.12%, respectively.

After calibrating the *SIDHRE-Q* model, the disease spread is observed with varying validated rapid antigen test performances and frequencies (Fig. 2). Sensitivity (the ratio of true positives to the total number of positives) and specificity (the ratio of true negatives to the total number of negatives) compared to gold-standard qRT-PCR were used as measures of test accuracy.

**Fig. 2.**
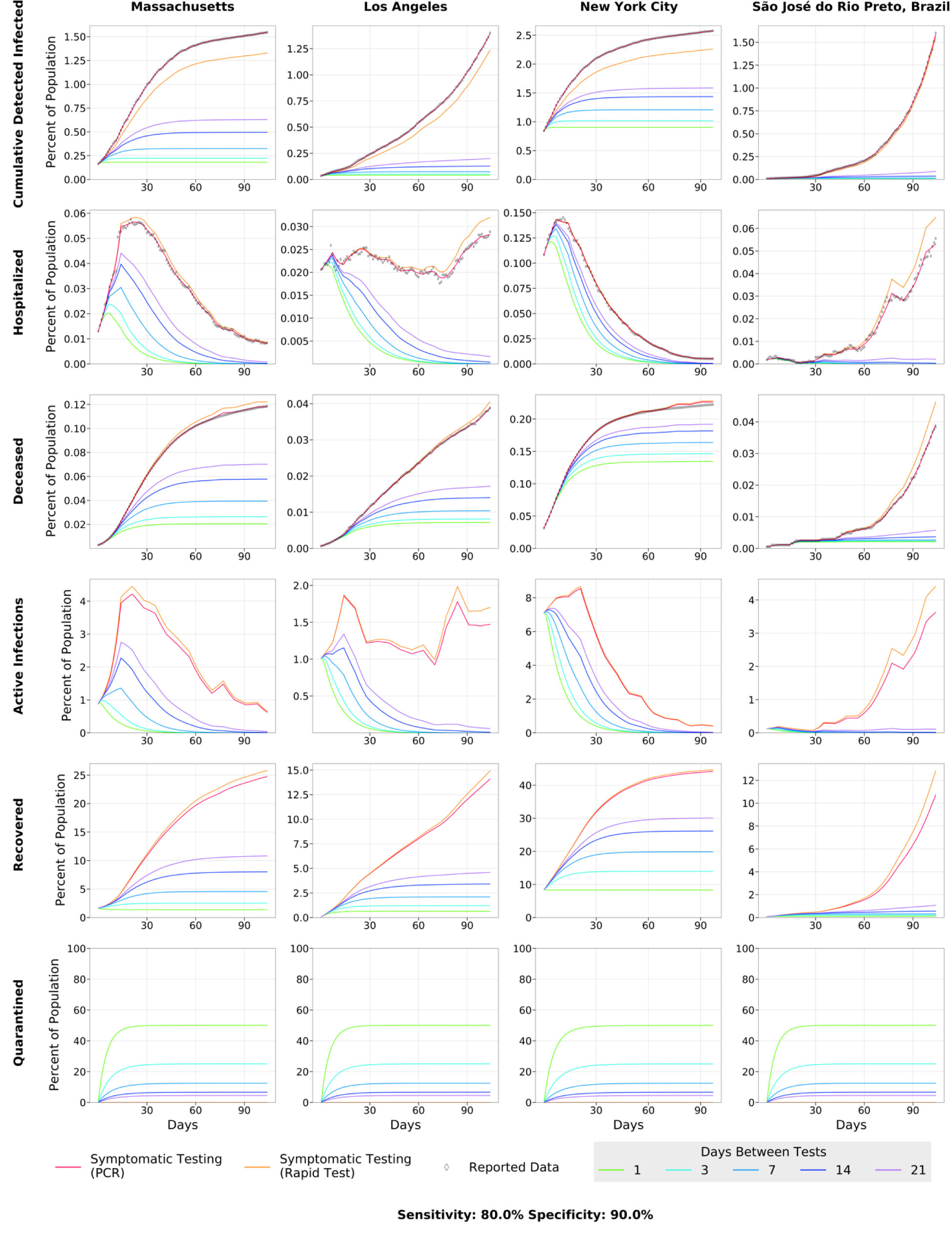
COVID-19 Outcomes in 3 US Regions and Brazil as a result of Frequent Rapid Testing Protocol using the *SIDHRE-Q* Model. The Cumulative Detected Infected, Hospitalized, Deceased, Active Infections, Recovered, and Quarantined are modeled over 105 days (top to bottom) using reported data from 4 global regions: Massachusetts, Los Angeles, New York City, and São José do Rio Preto in Brazil (left to right). The COVID-19 population spread and outcomes are modeled under a Rapid Testing Protocol (sensitivity 80%, specificity 90%) with variable testing frequencies ranging from 1-21 days between tests. This protocol is compared to a symptom-based Rapid Testing protocol and a symptom-based PCR protocol.

The rapid test frequency is varied while maintaining an accuracy of 80% sensitivity and 90% specificity, comparable to our clinical data collected in SJRP, Brazil. These testing scenarios are then compared to symptomatic testing, in which individuals receive a rapid test only when presenting symptoms, via either a rapid test or qRT-PCR. Since the primary testing regimen deployed in MA, LA, NYC and SJRP, Brazil is qRT-PCR-based and focused on symptomatic individuals, the symptomatic testing protocol via qRT-PCR is directly estimated from the data to be the rate *v* (Table 4).

The difference between the qRT-PCR and rapid test simulations (red and orange lines, respectively) is therefore only sensitivity of testing (Fig. 2). Test outcome probability in this model is a function only of whether an individual is infected and independent of other factors; one can consider this a lower bound on effectiveness of a strategy, as sensitivity and infectivity are often positively correlated with antigen testing. In this model with sensitivity *s* and frequency of testing *f*, the probability an individual is diagnosed in a testing window is given by the following:

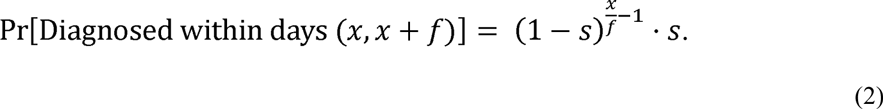

To better understand the effect of rapid testing frequency and performance on healthcare capacity and mortality rates, we simulate the testing strategy with 30%-90% sensitivity each with 80% or 90% specificity against the symptomatic testing strategy (Supplementary Fig. 3).

As per our hypothesis, frequency and symptom-based testing dramatically reduced infections, simultaneous hospitalizations, and total deaths when compared to the purely symptom-based testing regimens, and infections, hospitalization, and death were reduced as frequency increased. Although testing every day was clearly most effective, even testing every fourteen days with an imperfect test gave an improvement over symptomatic testing with qRT-PCR. While the strategy works best when implemented at the very beginning of an outbreak, as demonstrated by the results in SJRP, Brazil, it also works to curb an outbreak that is already large, as demonstrated by the results in NYC. The difference between frequencies is more noticeable when the testing strategy is applied to the outbreak in NYC, leading us to hypothesize that smaller outbreaks require a lower testing frequency than larger ones; note the difference between the dependence on frequency to curb a small initial outbreak in SJRP, Brazil versus a large one in NYC (Fig. 3).

**Fig. 3.**
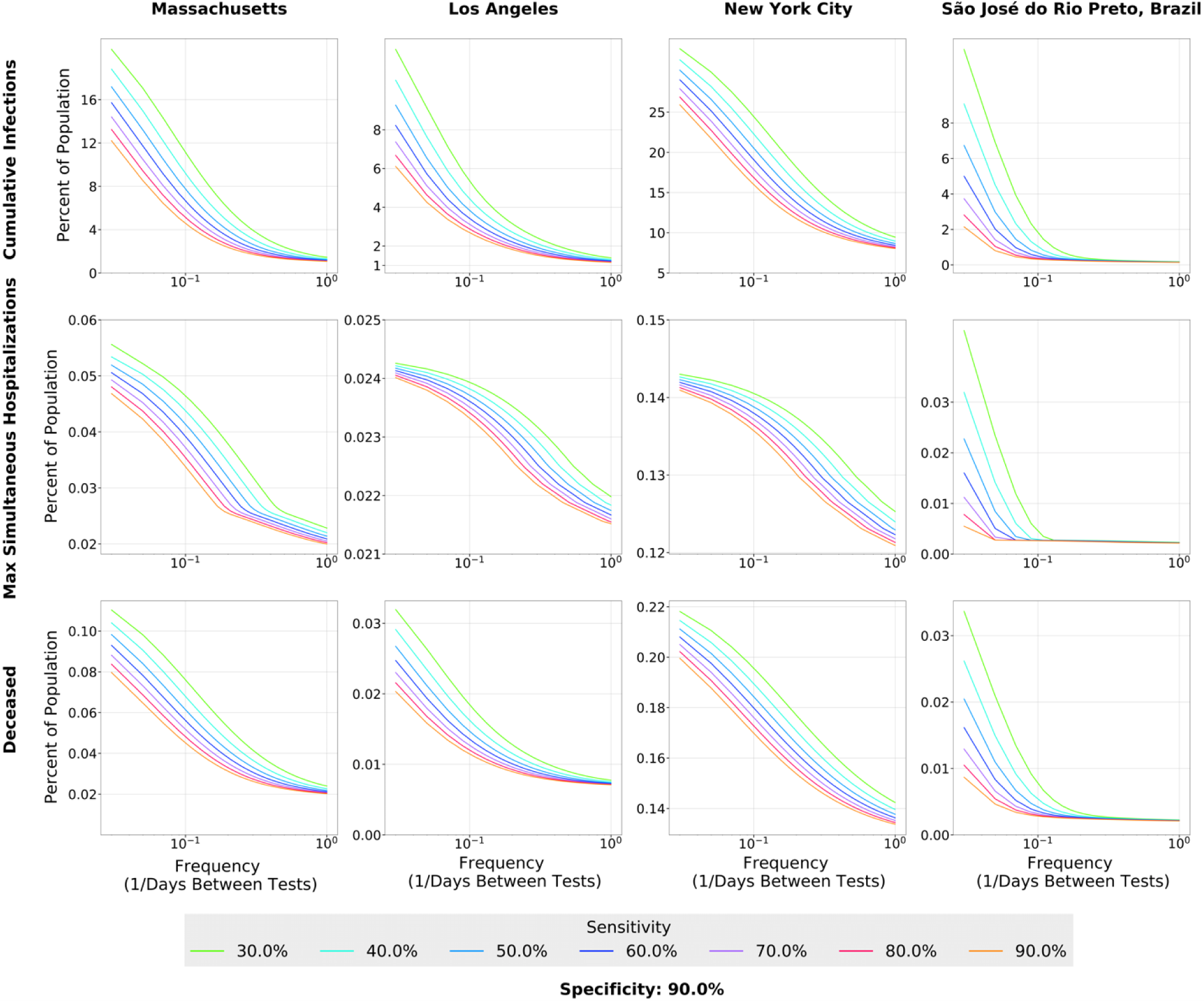
Effect of Rapid Testing Protocol under variable testing sensitivities (30%-90%) and increasing frequency under the *SIDHRE-Q* Model. The Cumulative Infections, Maximum Simultaneously Hospitalized, and Deceased populations are modeled for Massachusetts, Los Angeles, New York City, and São José do Rio Preto in Brazil with a 90% test specificity.

For test performance of 80% sensitivity and 90% specificity, the percent of the population that has been infected in total from the beginning of the outbreak to mid-July drops from 18% (MA), 11% (LA), 26% (NYC), and 11% (SJRP, Brazil) to 3%, 2%, 12%, and 0.26%, respectively, using a weekly rapid testing and quarantine strategy (with regards to predictions of overall infection rates, other studies based on seroprevalence and epidemiological predictions have reached similar conclusions ^49, 50^). If testing is increased to once every three days, these numbers drop further to 1.6% (MA), 1.4% (LA), 9.5% (NYC), and 0.19% (SJRP, Brazil) (Supplementary Table 1).

To further examine the relationship between frequency and sensitivity, we model the maximum number of individuals in a given state over the 105-day time period for four geographic regions (Fig. 3, Supplementary Fig. 4). In all four geographic regions, as frequency of testing increases, the total infections, maximum simultaneous hospitalizations, and total deaths converge to small percentages regardless of the sensitivity at high frequencies. For example, the predictions show that for the outbreak in LA, a testing strategy started on 1 April of every 10 days using a test of sensitivity 90% would have resulted in 2.5% of the population having been infected, while using a test of sensitivity 30% would require a strategy of every 5 days to achieve the same number. Thus, we conclude that frequency is more important than sensitivity to control the outbreak using a test-based strategy, and a large range of sensitivities prove effective when testing sufficiently often (Supplementary Fig. 4-5)^29, 51^. The following subsection contains a discussion of a location-based method for varying the exact frequency of testing based on evolving outbreaks. Frequency of testing can be significantly reduced to effectively contain the disease once the initial outbreak has been controlled; it is clear that this takes only a matter of weeks (Fig. 2).

On the other hand, according to the specificity of the rapid test and the quarantine duration, larger testing frequency result in a larger percent of the population quarantined (Fig. 2). Assuming a 90% rapid test specificity and 10-day quarantine duration, for the 1-, 3- and 7-day frequencies almost 48%, 24% and 12% of the population, respectively, would be quarantined. This figure may be reduced with additional rules for exiting quarantine early, such as after complementary testing. An example of such a strategy is that individuals who test positive are required to either quarantine for two weeks or produce two consecutive negative rapid tests in the two days following their positive result. Assuming 80% sensitivity and 90% specificity, those individuals will reenter the public while still infected with probability 0.04. If uninfected, that individual will exit quarantine after two days with probability 0.81. However, a compromise between the reduction of infections and the proportion of the population in quarantine would be part of the planning for the appropriate testing protocol in each community or region.

While high frequency may be necessary to contain a large outbreak initially, relatively infrequent testing, such as every one or two weeks, is sufficient to keep controlled outbreaks small, while reducing the number of quarantined individuals to less than 10% of the population using a two-week mandatory quarantine.

Additionally, quarantine adherence is of essential importance to the success of this strategy, and we assume near-perfect quarantine compliance with a small transmission rate due to diagnosed individuals (Table 4). Therefore, measures are needed to ensure quarantine is widely adhered to (Supplementary Fig. 8). Recent research has identified a number of ways to increase quarantine compliance, including compensating for wages lost, providing quarantine facilities and effective handling of the health crisis. ^52–55^

Additionally, while high frequency may be necessary to contain a large outbreak initially, relatively infrequent testing, such as every one or two weeks, is sufficient to keep controlled outbreaks small, while reducing the number of quarantined individuals to less than 10% of the population using a two-week mandatory quarantine.

### A County-Based Testing Strategy Offers a Cost-effective Approach to Large-scale COVID-19 Surveillance

To examine the effects of resource-strategic testing schemes, we modeled the COVID-19 prevalence by varying testing frequency across counties of California. For this analysis, only California was analyzed because of the accessibility of the county level data and the variability of spread dynamics of the outbreaks between counties. In this scheme, the percent of active infected detected individuals in a county determines the frequency of testing. We define thresholds for the number of active detected infections that, when hit, initiate testing protocols of different frequencies depending on the threshold hit. We first tested evenly spaced thresholds for the number of detected active infections up to 1% of the population, but later adopted thresholds that were determined according to Equation 3. In Equation 3, *D* = population of state **D** at the time of testing. *T* = number of active infections which, if reached, initiates everyday testing. The days between tests are rounded to the closest integer value.

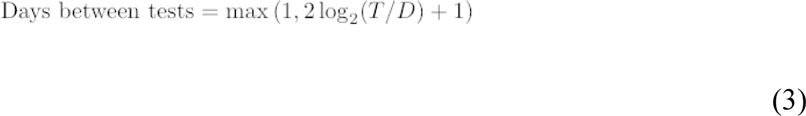

The days between tests are chosen such that the detected active infections should remain near to or below *T*. If the initial detected active infections are greater than *T*, then the testing frequency of 1 will cause infections to rapidly drop. Both the threshold at which everyday testing begins and the coefficient of *log_2_T/D* can be modified to produce a strategy that is more or less frequent in testing or resource effective; a range of days between tests from 14 days to 1 day are used (Fig. 4a).

**Fig. 4.**
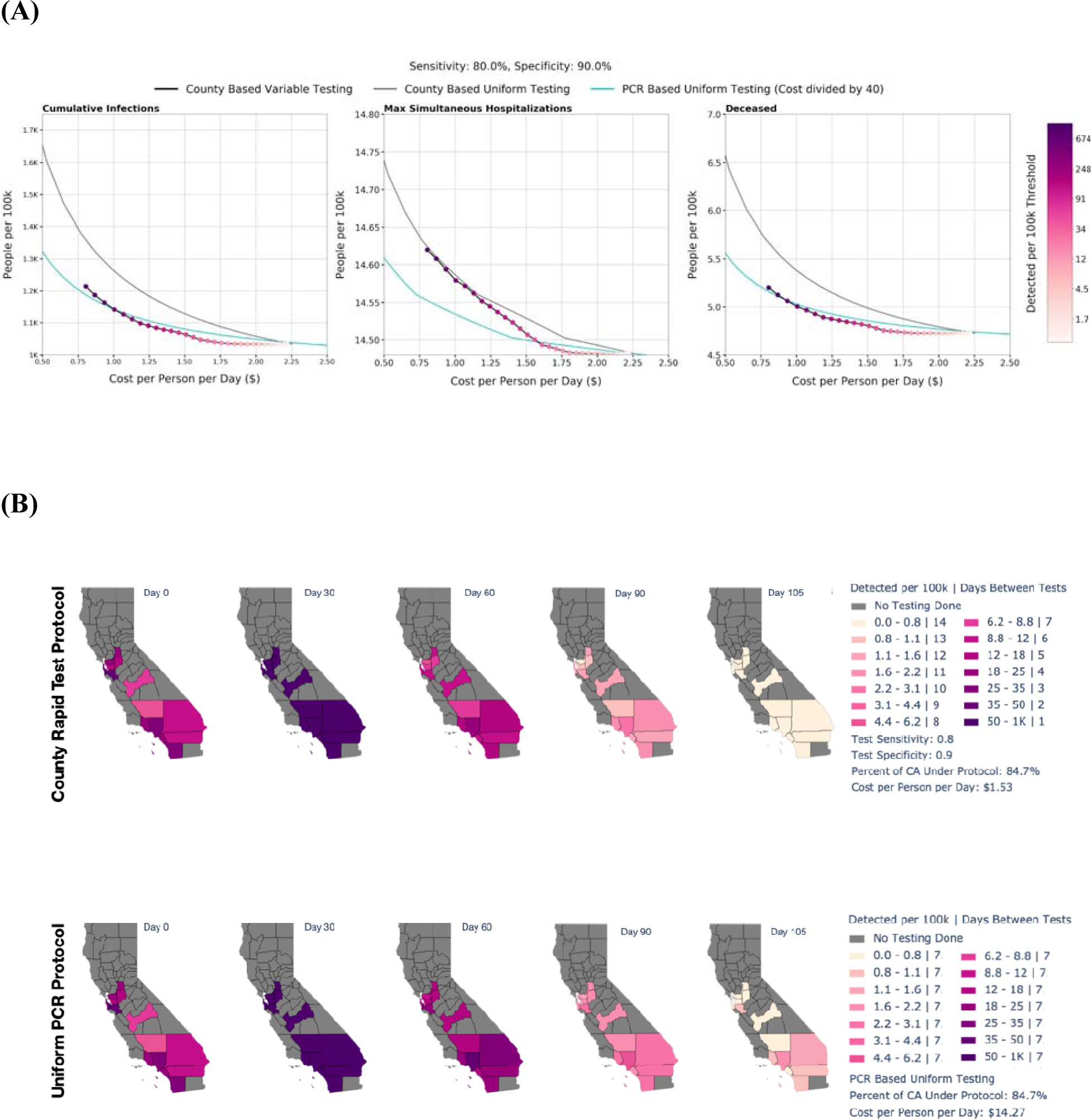
Effect of County Based Rapid Testing strategy on COVID-19 outcomes in California. This protocol varies testing frequency in accordance to the number of recorded cases; the threshold for number of active infections which, if reached, signals to commence everyday testing (the highest frequency considered). A Rapid Test with an 80% sensitivity and 90% sensitivity versus is used in this deployment strategy. Shown is the total cost per person per day versus the cumulative infections, maximum simultaneously hospitalized, and cumulative deaths with varied thresholds for all of CA is shown. The County Based Rapid Testing strategy is compared to uniform testing, which distributes the same number of total tests used in the county strategy, albeit evenly across each county. The effects of uniform testing are modeled for both a Rapid Testing protocol and a qRT-PCR protocol (A). The effects of County Based Rapid Test Protocol and Uniform PCR Protocol on active infected detected population over time in CA are shown (B). The legend denotes the thresholds at which testing frequency is determined, the testing frequencies, the percent of CA population under the strategy, and the cost per person per day.

The purpose of this strategy is to tailor testing based on the specific characteristics of unique outbreaks in different regions. A scan over different choices of *T* is shown in Fig. 4b; the threshold we choose in Fig. 4a is 0.05% because it is successful in curbing the outbreak in California within the time period we consider. Our analysis could be redone to select another effective fine-grained strategy in other states or regions. The cost analysis is based on cost per test - $7 per rapid test and $100 per PCR test - times number of tests used. Clearly that calculation neglects the costs of storing, distributing, and administering tests, as well as monitoring incoming results. The costs associated with these logistics would vary with differing policies dictating the use of rapid tests; significantly, whether they would be administered at home with self-reported results or in a testing facility or workplace for validation purposes. For example, a company may choose to use the rapid tests to scan employees before allowing them to enter the workplace, in a way similar to existing temperature checks. The cost of this particular application would be minimal beyond that of the actual tests. Such costs would inevitably be greater for PCR tests, which require a specialized testing facility, significant equipment, and highly trained personnel.

Using a rapid test with a sensitivity of 80% and a specificity of 90%, the county-based testing with threshold 0.05% reduces the active infections from 0.94% to 0.0005%, while the uniform strategy with tests administered every 7 days results in double the number of active infections (Fig. 4a). As the threshold is reduced, the total cost increases while the cumulative infections, maximum percentage hospitalized, and cumulative deaths all decrease (Fig. 4b). Appropriate choice of threshold is dependent on the severity of outbreaks in a specific region and available resources, both logistically and fiscally. With regional data, such as that from California used to produce Fig. 4b, this study can be reproduced to calculate an efficient testing strategy that will effectively curb outbreaks of differing severities in any geographic entity. This analysis does not include any delays in ramping testing up and down. If one were to reproduce this analysis for a given testing strategy, a fixed-time delay could be introduced, depending on the relevant logistical constraints.

Strategy B in Fig. 4 consists of qRT-PCR testing uniformly applied to the highlighted population with a frequency of once weekly. The average cost per person per day is just under $15. Despite this frequency and the accuracy of qRT-PCR, the strategy does not succeed in curbing the spread as fast as strategy A, which uses a testing sensitivity and specificity of 80% and 90%, respectively, and testing frequency that vary between counties depending on the proportion of their population that is currently infected. The total cost for strategy A is estimated at a fraction of the other at $1.53 per person per day.

## DISCUSSION

In this study we examine the potential effects of a novel testing strategy to limit the spread of SARS-CoV-2 utilizing rapid antigen test screening approaches. Our clinical data and *SIDHRE-Q* modeling system demonstrate that 1) frequent rapid testing even at a range of accuracies is effective at reducing COVID-19 spread, 2) rapid antigen tests are a viable source for this strategy and diagnose the most infectious individuals, and 3) strategic geographic-based testing can optimize disease control with the amount of available resources. The public has witnessed and experienced symptomatic individuals being denied testing due to shortages, and few testing structures for asymptomatic or mildly symptomatic individuals – a significant source of disease spread. Though several factors contributed to the stymied early response measures, such as lockdown and quarantine protocols and adherence, severe testing bottlenecks have been a significant culprit ^56–58^. Early control measures have been shown to decrease lives lost by several orders of magnitude^59^. These challenges, though exacerbated during the early months of the pandemic, remain at the forefront of the public health crises.

Diagnosis of SARS-CoV-2 infection by qRT-PCR is the current standard of care, yet remains expensive and requires a laboratory and experienced personnel for sample preparations and experimentation. The turnaround time for results can be up to 10 days, preventing people from either leaving quarantine if they are negative, or delaying critical care and infecting others if they are positive ^12^. This current testing scheme moreover yields incomplete surveillance data on which response efforts such as societal reopening and hospital management depend. Though qRT-PCR is considered the gold-standard diagnostic method because of its high sensitivity and specificity, the logistical hurdles render it unrealistic for large-scale screening.

As qRT-PCR remains impractical for this strategy, and rapid tests are facing regulatory challenges because they do not perform with qRT-PCR-like accuracy, rapid test screening is either nonexistent in several countries or symptom-based. Even under best-case assumptions, findings have shown that symptom and risk-based screening strategies miss more than half of the infected individuals ^60^. Some have argued that the need for widespread testing is overstated due to the variability in test sensitivity and specificity ^61^. Here, we present alternative large-scale diagnostic tools to qRT-PCR, and show that test performance, though valuable, is secondary to widespread test frequency, which is enabled by accessibility and turnaround time. Furthermore, test affordability is essential for the successful implementation in communities most affected by infection and will to speed up the safe opening and functioning of the viral sectors of the economy.

Giordano et al. has modeled the evolution of SARS-CoV-2 spread, introducing a diagnosed state to elucidate the importance of population-wide testing ^29^. Larremore et al. has examined how various test sensitivities and frequencies affect the reproductive number ^21^. We build upon these findings to show how in affected United States and Brazil regions, population-wide frequent and rapid testing schemes, with sensitivities ranging from 30%-90%, can be more effective in curbing the pandemic than a PCR-based scheme. Integrating real-world surveillance and clinical data into our modeling system has allowed us to incorporate regional differences - such as variances in healthcare access, state health policy and adherence, state GDP, and environmental factors - under the same model. Significantly, our findings hold true across Massachusetts, New York City, Los Angeles, and São José do Rio Preto, Brazil. We also present the economic considerations of these testing regimes, showing that widespread rapid testing is more cost efficient than less frequent qRT-PCR testing. In line with these economic considerations, our model demonstrates the effectiveness of a geographic-based frequent testing regime, in which high disease prevalence areas receive more frequent testing than low disease prevalence areas.

Since COVID-19 is known to affect certain demographics differently, modeling would benefit from incorporating demographic information correlated with disease progression and spread to define sub-models and sets of parameters accordingly. Age, pre-existing conditions, job types, and density of population are examples of possible categories, each of which influence the risk of contracting and/or dying from COVID-19. Further studies would benefit from incorporating these ideas to better understand the effectiveness of rapid testing on identifying potential super spreading events. Future public health prevention programs should use the proposed modeling system to develop and test scenarios for precision testing and prevention.

Our findings also point to low-cost tools for implementation of this testing strategy, such as a rapid antigen-based test for the detection of SARS-CoV-2 proteins. We show that the rapid antigen tests perform with a range of accuracies under which disease spread can be dramatically mitigated under our model. Notably, the sensitivity is correlated to the individual’s viral load, effectively diagnosing those who are potentially the most infectious with the highest accuracy. Our findings are significant because rapid antigen tests are cheaper than qRT-PCR, can be mass produced to millions per day, present results within 15 minutes, and can be administered by a nonexpert without a lab or special equipment.

There are several policy implications for these findings. First, our model supports that systems of high frequency rapid testing should be implemented as a first-line screening method. This can be first enabled by a more holistic regulatory evaluation of rapid diagnostics, such that policy emphasizes accessibility and turnaround time even under a range of accuracies. One can imagine a less accurate, though rapid method of first-line screening in schools, public transportation, and airports, or even at home, and a qRT-PCR-based method for second-line screening (testing those who present severe symptoms or have been in contact with infected individuals, testing in a clinical setting, etc). At home tests require a built-in digital reporting capability^62^; rapid antigen test results can be sent to local health centers with reciprocal instructions regarding updated test frequency guidelines to enable adaptive testing strategies.

Second, our cost analysis and rapid antigen test data present a viable and potentially more cost-effective method for screening. Third, our county-based testing scheme presents a possible method for wide-scale screening while optimizing resources. Future studies should investigate how this selective testing strategy can be applied to different location scales to further inform health policy. Moreover, though our models analyze regions in the United States and Brazil, similar testing strategies can be considered globally in both resource limited and abundant settings due to the greater accessibility of rapid tests compared to qRT-PCR. This model can be further tailored to the pandemic course as we gain further evidence regarding SARS-CoV-2 re-infection rates.

We emphasize that integral to the effectiveness of diagnostic schemes is 1) the proper adherence to quarantine and public health measures and 2) the combined use of a variety of diagnostic methods including nucleic acid, antigen, and antibody tests. According to these models, rapid antigen tests are an ideal tool for first-line screening. Clinical molecular tests such as qRT-PCR are vital to the diagnostic landscape, particularly to re-test suspected cases that were negative on the rapid test. Because rapid tests present a higher rate of false negatives, methods such as qRT-PCR remain integral to second-line screening. Antibody tests provide important information for immunity and vaccination purposes as well as epidemiological surveillance. This model also assumes that individuals will quarantine themselves before being tested and for 10 days following a positive diagnostic result and will not be infected while waiting for the qRT-PCR results. It is important to acknowledge the working definition of quarantine. The states containing quarantined individuals (**Q_U_, Q_R_** and **D**) are defined as consisting of a population that is meant to be quarantined, not a population that is necessarily in perfect compliance with the mandate that they remain fully isolated from the population. Quarantine is assumed to be imperfectly executed and the model accounts for a small, tunable interaction between quarantined states and the general population, hence it is conservative.

There are important limitations to be considered in this model. Differences in disease reporting between the geographical regions and the incomplete nature of COVID-19 surveillance data, often due to the lack of testing or delays in reporting, are not considered in the model. It is imperative that the testing results, hospitalization and death statistics, and changes in protocol are reported in real-time to scientists and policy makers so that models can be accurately tuned as the pandemic develops. Moreover, delays required to ramp testing strategies up or down are not considered. Infectivity variations between individuals is also not applied to this model, and future clinical studies should gather data on asymptomatic presenting COVID-19 cases. Non-compliant quarantine behaviors and possible infections during testing waiting times are also not included in the calculations. The model also does not take into account infrastructural limitations, such as hospital capacity and testing space, which depend on factors beyond the scope of this analysis. Though the rapid antigen test offers several advantages such as affordability, fast turnaround time, and ease of mass production, we are assuming that there are systems in place to implement frequent and safe low-cost screening across different communities and settings.

Our model underscores the need for a point-of-care or at-home test for frequent screening, particularly as lockdown restrictions ease. Regulatory agencies can work towards evaluating rapid tests to alternative standards other than comparison to high sensitivity molecular diagnostics, as our model shows that frequency and scale of testing may overcome lower sensitivities. Rather, we can refocus policy to implement first-line screening that optimizes accuracy with efficiency and equitability.

## METHODS

### Development of Direct Antigen Rapid Tests for the Detection of SARS-CoV-2

We developed a direct antigen rapid test for the detection of the nucleocapsid protein or spike glycoprotein from SARS-CoV-2 in nasal or nasopharyngeal swab specimens as previously described ^63^. Briefly, the rapid antigen tests are immunochromatographic format with a visual readout using anti-N or anti-S mouse monoclonal antibodies (E25Bio, Inc., Cambridge, MA, USA) that are either coupled to 40 nm gold nanoparticles (Abcam, Cambridge, UK) or adsorbed to nitrocellulose membranes (Sartorius, Goettingen, Germany). Each rapid antigen test has a control area adjacent to the paper absorbent pad; the control is an anti-mouse Fc domain antibody (Leinco Technologies, Fenton, MO, USA) that will capture any of the antibody-conjugated gold nanoparticles to generate a control visual signal. A visual signal at the test area reflects SARS-CoV-2 N or S that is “sandwiched” between an anti-N or anti-S antibody adsorbed to the nitrocellulose membrane and a second anti-N or anti-S antibody covalently coupled to visible gold nanoparticles.

### Validation of Direct Antigen Rapid Test for the Detection of SARS-CoV-2

In a retrospective study of nasal swab specimens form human patients, we compared the accuracy of the rapid antigen test for detection of SARS-CoV-2 N to the viral loads of individuals. All individuals were symptomatic between 1-10 days of fever. Nasal swab specimens (n=158) were tested following approved human subjects use protocols. The nasal swab specimens were banked frozen from suspected patients submitted to PATH for routine COVID diagnosis. Prior to using the rapid test, the nasal swab specimens were validated by qRT-PCR using the FDA EUA ThermoFisher/AppliedBiosystems TaqPATH COVID-19 Combo Kit (ThermoFisher, Waltham, MA USA). The primary study under which the samples and data were collected received ethical clearance from the PATH Research Ethics Committee, protocol number 00004244; all participants provided written informed consent for the use of the samples. The nasal swab specimens were de-identified, containing no demographic data, prior to analysis, and the experiments were performed in accordance with relevant guidelines and regulations.

The nasal swabs were originally collected in 1 mL PBS, where 50 μl was mixed with 50 μl of Solution Buffer (0.9% NaCl and 0.1% Triton X-100). The 100 μl mixture was then pipetted onto the rapid antigen test for SARS-CoV-2 N detection and allowed to react for 15 minutes. After processing of the rapid antigen test, the visual positive or negative signal was documented.

Additionally, in a retrospective study of nasopharyngeal swab specimens from human patients, we compared the accuracy of the rapid antigen test to the viral load of individuals. Nasopharyngeal swab specimens (n = 121) were tested in Brazil following approved human subjects use protocols. The age of study participants ranged from 1 to 95 years with an overall median of 37 years (interquartile range, 27–51 years), and 62% were female. All individuals were symptomatic between 1-10 days of fever. The demographic summary of the patients are included in Supplementary Table 2. The nasopharyngeal swab specimens were banked refrigerated or frozen samples from suspected patients submitted to the lab for routine COVID diagnosis. Prior to using the rapid test, the nasopharyngeal swab samples were validated by qRT-PCR using GeneFinder^TM^ COVID-19 Plus Real*Amp* Kit (OSANGHealtcare, Anyang-si, Gyeonggi-do, Republic of Korea I). The primary study under which the samples and data were collected received ethical clearance from the Faculdade de Medicina de São José do Rio Preto (FAMERP), protocol number 31588920.0.0000.5415; all participants provided written informed consent for the use of the samples. All excess samples and corresponding data were banked and de-identified prior to the analyses, and the experiments were performed in accordance with relevant guidelines and regulations.

Nasopharyngeal swab specimens (1 mL) were concentrated using Vivaspin 500 centrifugal concentrators (Sartorius, Goettingen, Germany) at 12,000 x g for 10 minutes. The concentrated nasopharyngeal swab specimen retentate was transferred to a collection tube and the rapid antigen test for SARS-CoV-2 spike detection was inserted into the tube with the retentate and allowed to react for 15 minutes. After processing of the rapid antigen test, the visual positive or negative signal was documented.

Both nasal and nasopharyngeal swabs were used for the detection of SARS-CoV-2 N and S, respectively. Some studies have shown higher efficacy of nasopharyngeal swabs for PCR tests; the similar results between our two cohorts are likely due to the different proteins being detected^64, 65^.

### Data for Modeling

As of August 2020, the United States and Brazil have the highest number of confirmed COVID-19 cases and deaths worldwide, with both countries reporting their first case on 26 February 202) ^1^. Although several affected US regions could have been modeled, we look at data from Massachusetts, New York, and Los Angeles: these regions each contained “hotspots”, or areas of surging COVID-19 cases, at different points in time during the pandemic and have publicly available government-provided surveillance data. Our model is fit using data over 105 days beginning on April 1 for Fig. 2 and Fig. 3, and 105 days beginning on April 10 for Fig. 4 (see “Modeling Parameters” in Methods). In order to understand the various testing proposals on a global scale, we performed our clinical study in and expanded the modeling study to Brazil. The specific data we use to fit our model are cumulative confirmed cases, total deaths, and number of daily hospitalizations due to COVID-19. This surveillance data was retrieved from government-provided online databases ^66–72^.

### Modeling Parameters

Equation 1 below provides the exact differential equations governing the model.

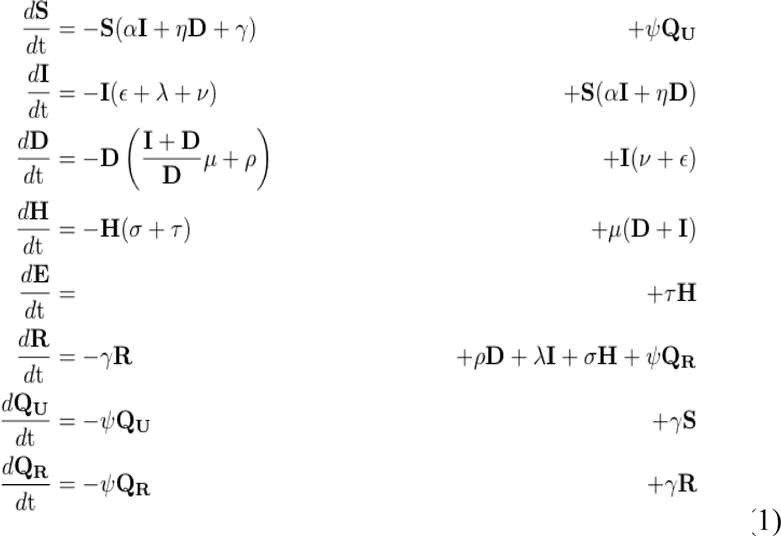

From Table 4 describing each parameter, note that each of their values are the inverse of the average rates at which a transition is made. For example, the term is set equal to the population exiting quarantine per day, with days. The assumption made is that the distribution of time already spent in quarantine is approximately uniform among the quarantined population at any given time. The uniform rate approximation breaks down during periods when flows between states are changing rapidly within a matter of days, such as in early stages of the pandemic. The result of running the model with fixed infection and quarantine times as well as a discussion of how that change is incorporated in Supplementary Fig. 9. The mean value method of assigning values to parameters is standard in epidemiological modeling ^29, 73^.

In order to determine the numerical values of the parameters defining the flows between states, we use a least squares regression to find fits for each seven day interval. All data points within each interval and from each data set are fit collectively within each interval (the resulting fits do not represent the mean of separately calculated fits). This procedure allows the model to take into account the time dependent nature of the parameters, which rely on factors such as social distancing regulations and changes in testing capacity. We also fit window sizes between 1 and 21 days and find that while the fit degrades with larger window size, the overall shape of the curves do not change. We choose seven days assuming policy changes take a week to become effective and that reasonably parameters can be expected to change within this time period without causing problems with overfitting. Also, the seven day window size accounts for the fact that often data is not reported as diligently over the weekend. Time series of the values of the parameters for the geographic locations discussed in this paper are included in Supplementary Fig. 6.

Given the restrictions on data available for the populations of various states, varying all of the parameters results in an over parameterized system. Therefore, a subset of the model parameters are fit while the others are either extracted from other sources; see Table 4. The fitting procedure minimizes the sum of the squared residuals of the normalized total cases, current daily hospitalizations, cumulative deaths, and percentage of total infected individuals currently hospitalized. The first three are present in the data sets while the latter is derived from the estimates of the ratio between infected undetected to infected detected individuals from the CDC Laboratory Seroprevalence Survey Data ^74^. Each of these data sets are normalized to maintain equal weight for least squares optimization.

While this ratio changes over time, the percentage of infected individuals developing severe symptoms should remain roughly constant throughout the course of the epidemic in the different locations studied.

We consider the data sets for outbreaks in MA, NYC, LA, and SJRP, Brazil ^66–71^. While each location has testing and fatality information dating back to January, hospitalization data was not included until late March (for NYC and SJRP) and April (for MA and LA). Hence we begin our fitting procedure and testing strategy on 1 April for each of the data sets; by this point, the outbreak is advanced in NYC, substantial in MA, non-negligible, but far from its peak, in LA, and in early stages in SJRP, Brazil. Starting simulations at various stages of the outbreak allows one to see the difference in results between when a testing strategy is administered.

In order to determine the effectiveness of the county-based strategy when applied to the state of California, we also fit all of the counties in California with a population greater than 1.5% of that of the entire state and with greater than zero deaths. The results do not depend on these selections, but instead suggest a practical criteria to administer limited resources. The fitting is done starting 10 April for these counties, as at this point the outbreak is sufficiently well-documented in each to successfully model. For the county-level data we compute a seven day running average of each of the data sets to which we then fit in order to smooth out fluctuations in the data, likely due to reporting, which are more significant here than in the other data sets considered, as the county populations are smaller and hence discrepancies impact the smoothness of the data more. The fits for each of the counties can be found in Supplementary Fig. 7.

As one can see from Fig. 1, these data sets are particularly not smooth, which indicates inefficiencies in reporting. Additionally, it is difficult to gauge their consistency within the dates provided or to compare between locations, as reporting mechanisms changed over time within the same locations. Despite this lack of consistency, our model and fitting mechanism was successful in reproducing the progress of the outbreak in each data set studied.

## Data Availability

The authors confirm that the data supporting the findings of this study are available within the article and/or its supplementary materials; any other data will be made available upon request. Our code can be found on github: https://github.com/badeaa3/COVID19_Rapid_Testing.

## DATA AVAILABILITY

The authors confirm that the data supporting the findings of this study are available within the article and/or its supplementary materials; any other data will be made available upon request.

## CODE AVAILABILITY

Full code can be found on github: https://github.com/badeaa3/COVID19_Rapid_Testing. The code is written using python with the packages scipy, numpy, lmfit, matplotlib and plotly ^75–79^.

## Acknowledgments

**General**: We thank Professor Lee Gehrke for critical reading of the manuscript.

**Funding:** EN is funded by Tufts University DISC Seed Grant. MLN is supported by a FAPESP grant (#2020/04836-0) and is a CNPq Research Fellow. AFV is supported by a FAPESP Fellow grant (#18/17647-0). GRFC is supported by a FAPESP Fellow grant (#20/07419-0). BHGAM is supported by a FAPESP Scholarship (#19/06572-2). The funders had no role in the design of the study; in the collection, analyses, or interpretation of data; in the writing of the manuscript, or in the decision to publish the results.

**Author contributions:** Conceptualization: BBH. Formal analysis: BN, AB, AR, MB, NS, ARG, AV, GCDS, TMILDS, BHGAM, MMM, GRFC, FQ, AFNR, MLN, ENN, IB, BBH. Funding acquisition: IB, BBH. Investigation: BN, AB, AR, MB, NS, ARG, AV, GCDS, TMILDS, BHGAM, MMM, GRFC, FQ, AFNR, MLN, ENN, IB, BBH. Methodology: BN, AB, AR, MB, NS, ARG, AV, GCDS, TMILDS, BHGAM, MMM, GRFC, FQ, AFNR, MLN, ENN, IB, BBH. Project administration: MLN, IB, BBH. Resources: MLN, IB, BBH. Supervision: MB, MLN, ENN, IB, BBH. Validation: BN, AB, AR, MB, ENN, BBH. Visualization: BN, AB, AR, MB, AV, ENN, BBH. Writing— original draft: AR, BBH. Writing—review and editing: BN, AB, AR, MB, NS, ARG, AV, GCDS, TMILDS, BHGAM, MMM, GRFC, FQ, AFNR, MLN, ENN, IB, BBH.

**Competing interests:** BN, AB, AR, MB, NS, AG, and BBH are employed by E25Bio Inc. (www.e25bio.com), a company that develops diagnostics for epidemic viruses. BBH and IB are co-founders of E25Bio. AV, GCDS, TMILDS, BHGAM, MMM, GRFC, FQ, AFNR, MLN, and ENN do not have any competing interests.

