## Supplementary Material for "Validating and modeling the impact of high-frequency rapid antigen screening on COVID-19 spread and outcomes"

**Supplementary Materials**

**Supplementary Table 1. Summary of results of COVID-19 outcomes in 3 US Regions and Brazil as a result of Frequent Rapid Testing Protocol using SIDHRE-Q Model.**

**Supplementary Table 2. Demographic and clinical summary of patients evaluated by DART for detection of SARS-CoV-2 spike glycoprotein.**

**Supplementary Fig. 1. Performance of direct antigen rapid test (DART) for the detection of SARS-CoV-2 (A) nucleocapsid protein and (B) spike glycoprotein.** Shown are the percentile positive cases of the total positive population conditioned to qRT-PCR Cycle Threshold (Ct).

Percentile Positive ranks the samples in order of high Ct to low Ct. DART sensitivity is determined by calculating true positive agreement to qRT-PCR; the plot uses an ax^b^+c fit and 95% confidence intervals for the sensitivity.

**Supplementary Fig. 2. Graphical scheme displaying the relationships between the stages of quarantine and infection in *SIDHRE-Q* model:** **Q-U**, quarantine uninfected; **S**, susceptible (uninfected); **I**, infected undetected (pre-testing and infected); **D**, infected detected (infection diagnosis through testing); **H**, hospitalized (infected with life threatening symptom progression); **R**, recovered (healed); **E**, extinct (dead); and **Q-R**, quarantine recovered (healed but in quarantine by false positive testing).

**Supplementary Fig. 3.** **COVID-19 Outcomes as a result of Frequent Rapid Testing Protocol with variable test performances using *SIDHRE-Q* Model.** The Cumulative Detected Infected, Hospitalized, Deceased, Active Infections, Recovered, and Quarantined are modeled over 105 days (top to bottom) using reported data from 4 global regions: Massachusetts, Los Angeles, New York City, and São José do Rio Preto in Brazil (left to right). The COVID-19 population spread and outcomes are modeled under a Rapid Testing Protocol with variable testing frequencies ranging from 1-21 days between tests, and variable test performances: 90% specificity with 90% sensitivity (A), 70% sensitivity (B), 50% sensitivity (C), and 30% sensitivity (D); and 80% specificity with 90% sensitivity (E), 70% sensitivity (F), 50% sensitivity (G), and 30% sensitivity (H). This protocol is compared to a symptom-based Rapid Testing protocol and a symptom-based qRT-PCR protocol.

**Supplementary Fig. 4. Effect of Rapid Testing Protocol under variable testing sensitivities and increasing frequency under the *SIDHRE-Q* Model.** The Cumulative Infections, Maximum Simultaneously Hospitalized, and Deceased populations are modeled for Massachusetts, Los Angeles, New York City, and São José do Rio Preto in Brazil. The effect of increasing frequency of testing is modeled for various testing sensitivities (30%-90%) with an 80% specificity.

**Supplementary Fig. 5.** Missed infections - number of infections that were never diagnosed by the rapid test as a function of log(frequency) for a range of sensitivities with a 90% specificity. Models are shown for MA, LA, NYC, and SJRP.

**Supplementary Fig. 6**. **Time series of the four fitted parameters 𝛼, 𝜈, 𝜇, and 𝜏 (left to right) for MA, LA, NYC, and SJRP (top to bottom).** See Table 4 in the Methods section for an explanation of the parameters. The values are extracted every seven days from data provided by the respective regions. The parameters vary significantly over time and location. Flat points occur during the seven day windows where the parameters are held constant. The fitting procedure is also outlined in the Methods section.

**Supplementary Fig. 7.** **Time series of the three fitted pieces of data Cumulative Cases, Daily Hospitalized, and Cumulative Deaths (left to right) for each county receiving testing in CA;** Ventura (A), Stanislaus (B), Santa Clara (C), San Joaquin (D), San Francisco (E), San Diego (F), San Bernardino (G), Sacramento (H), Orange (I), Los Angeles (J), Kern (K), Fresno (L), Alameda (M). The counties included satisfy two requirements: population greater than 1.5% of the total CA population and nonzero total number of deaths at each point in time. The fitting procedure is outlined in the Methods section.

**Supplementary Fig. 8.** Dependence of total infections and deaths over the 105 day period in Massachusetts shown as a function of  $\eta$, the value of which indicates quarantine effectiveness, with $\eta$ =0 reflecting full compliance (no transmission due to quarantined individuals) and $\eta$ =1 reflecting no compliance (same transmission due to quarantined individuals as those not quarantined).

**Supplementary Fig. 9.** Comparison of using a mean value approximation as opposed to fixed duration quarantine and infection periods.  To test the validity of the mean value approximation, we repeat the simulations of the model but replace the single **I** state with 10 substates, each of which corresponds to a different day of infectivity, **D** with 10*10 sub-states, one for each (day infected, day diagnosed) combination, as well as the **Q** state with 10 sub-states corresponding to each day of quarantine.  From sub-state *n < 10* of **I**, there is a flow of value 1 to sub-state *n+1* of **I** as well as a flow into sub-state *n* of **D,** corresponding to rate of diagnosis.  The fixed duration model introduces more complexity than the mean value scheme, which is standard in epidemiological studies, and produces only minimally different results when simulated using otherwise identical models.

**Supplementary Tables**

**Supplementary Table 1. Summary of results of COVID-19 outcomes in 3 US Regions and Brazil as a result of Frequent Rapid Testing Protocol using SIDHRE-Q Model.**

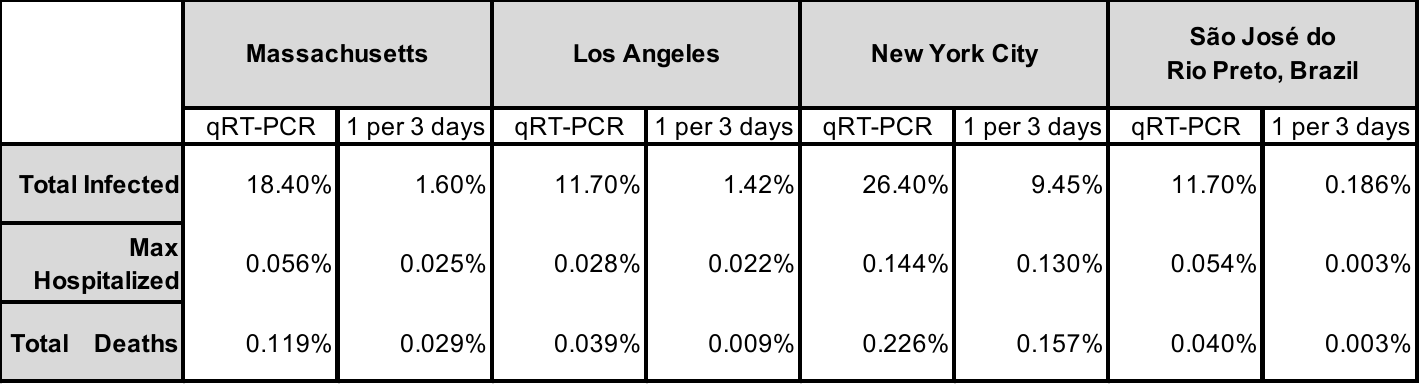

**Supplementary Table 2. Demographic and clinical summary of patients evaluated by the SARS-CoV-2 Direct Antigen Rapid Test (DART).** N response, N or mean of positive, and % or standard deviation for each group is presented. All samples (n=121) collected and tested in São José do Rio Preto, Brazil.

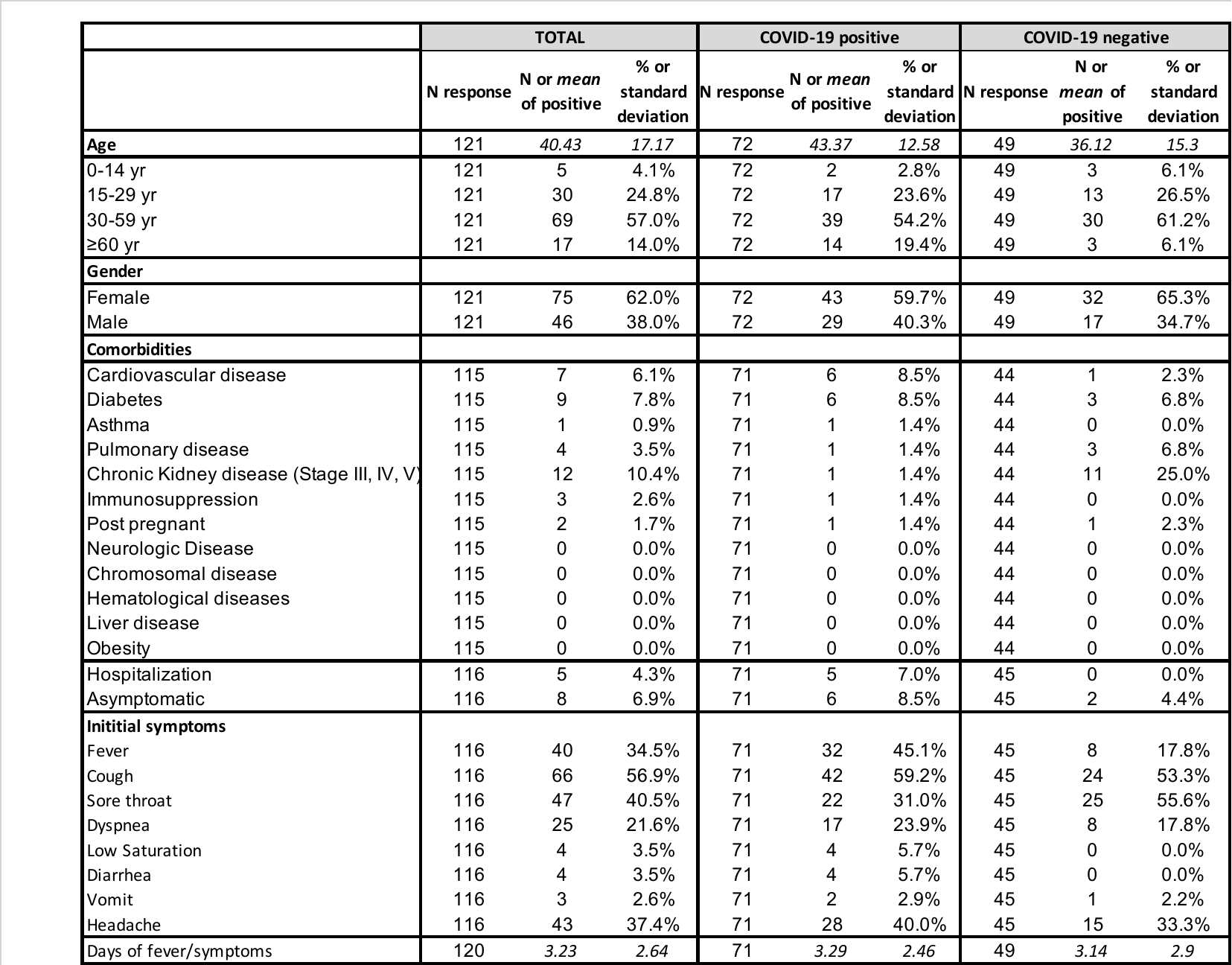

**Supplementary Figures**

**Supplementary Fig. 1. Performance of direct antigen rapid test (DART) for the detection of SARS-CoV-2 (A) nucleocapsid protein (n=158) and (B) spike glycoprotein (n = 121).** Shown are the percentile positive cases of the total positive population conditioned to qRT-PCR Cycle Threshold (Ct). Percentile Positive ranks the samples in order of high Ct to low Ct. DART sensitivity is determined by calculating true positive agreement to qRT-PCR; the plot uses an ax^b^+c fit and 95% confidence intervals for the sensitivity.

**(A)**
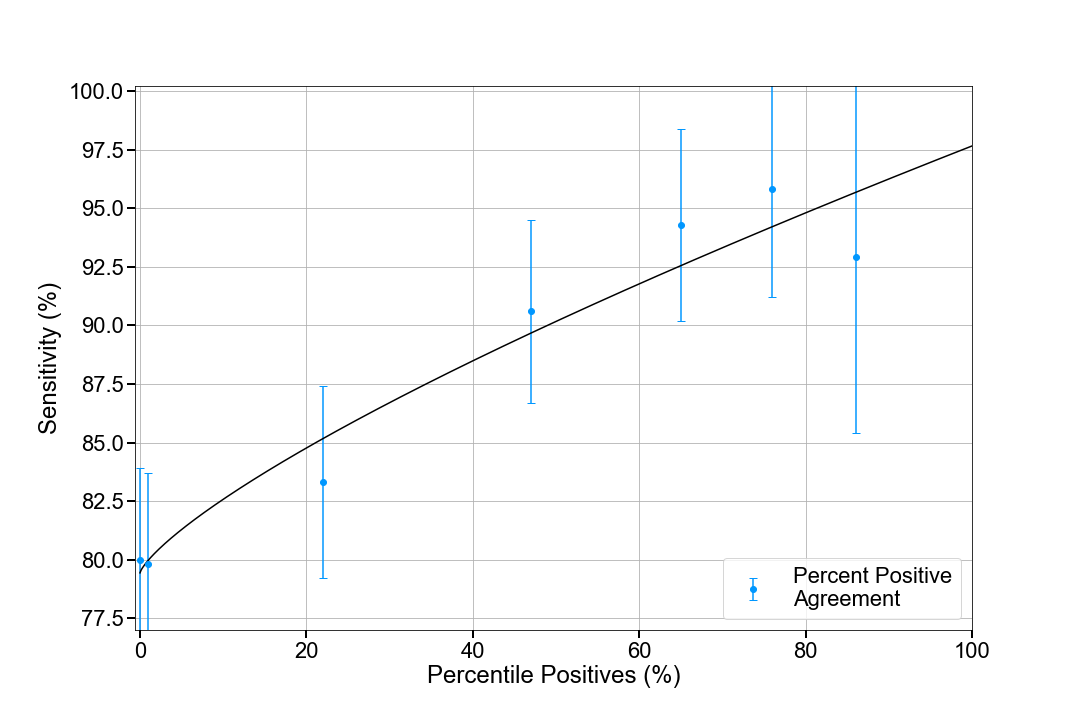

**(B)**
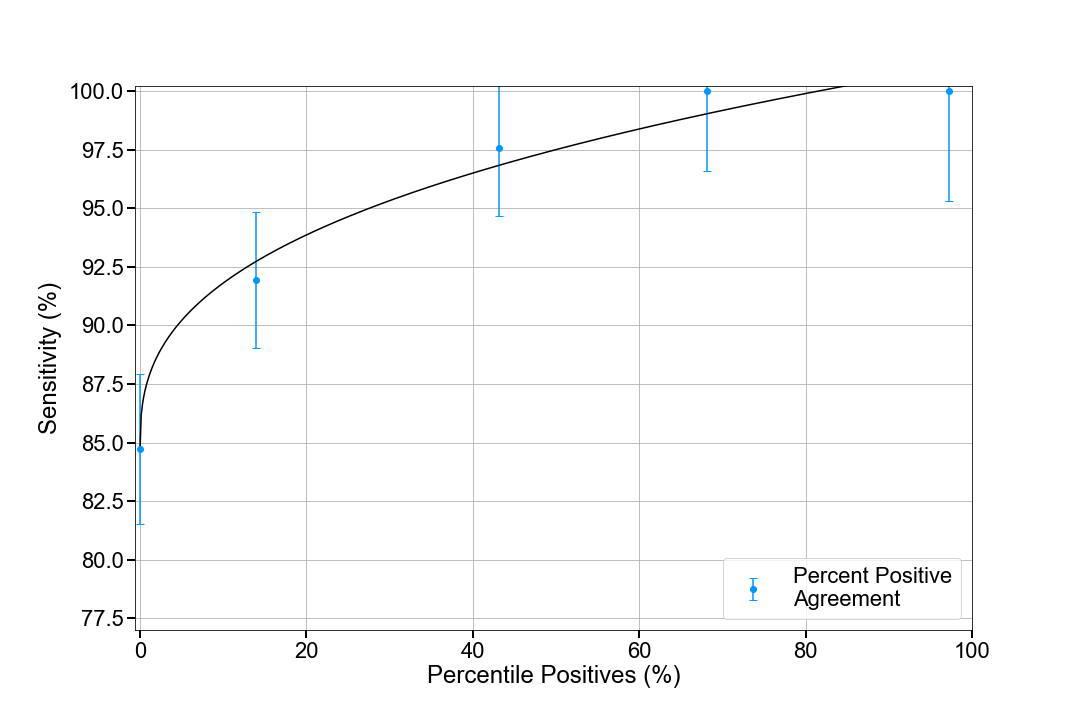

**Supplementary Figure 2. Graphical scheme displaying the relationships between the stages of quarantine and infection in *SIDHRE-Q* model:** **Q-U**, quarantine uninfected; **S**, susceptible (uninfected); **I**, infected undetected (pre-testing and infected); **D**, infected detected (infection diagnosis through testing); **H**, hospitalized (infected with life threatening symptom progression); **R**, recovered (healed); **E**, extinct (dead); and **Q-R**, quarantine recovered (healed but in quarantine by false positive testing).

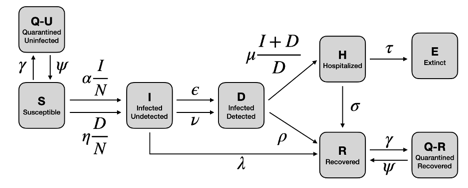

**Supplementary Figure 3.** **COVID-19 Outcomes as a result of Frequent Rapid Testing Protocol with variable test performances using *SIDHRE-Q* Model.** The Cumulative Detected Infected, Hospitalized, Deceased, Active Infections, Recovered, and Quarantined are modeled over 105 days (top to bottom) using reported data from 4 global regions: Massachusetts, Los Angeles, New York City, and São José do Rio Preto in Brazil (left to right). The COVID-19 population spread and outcomes are modeled under a Rapid Testing Protocol with variable testing frequencies ranging from 1-21 days between tests, and variable test performances: 90% specificity with 90% sensitivity (A), 70% sensitivity (B), 50% sensitivity (C), and 30% sensitivity (D); and 80% specificity with 90% sensitivity (E), 70% sensitivity (F), 50% sensitivity (G), and 30% sensitivity (H). This protocol is compared to a symptom-based Rapid Testing protocol and a symptom-based qRT-PCR protocol.

**(A)**

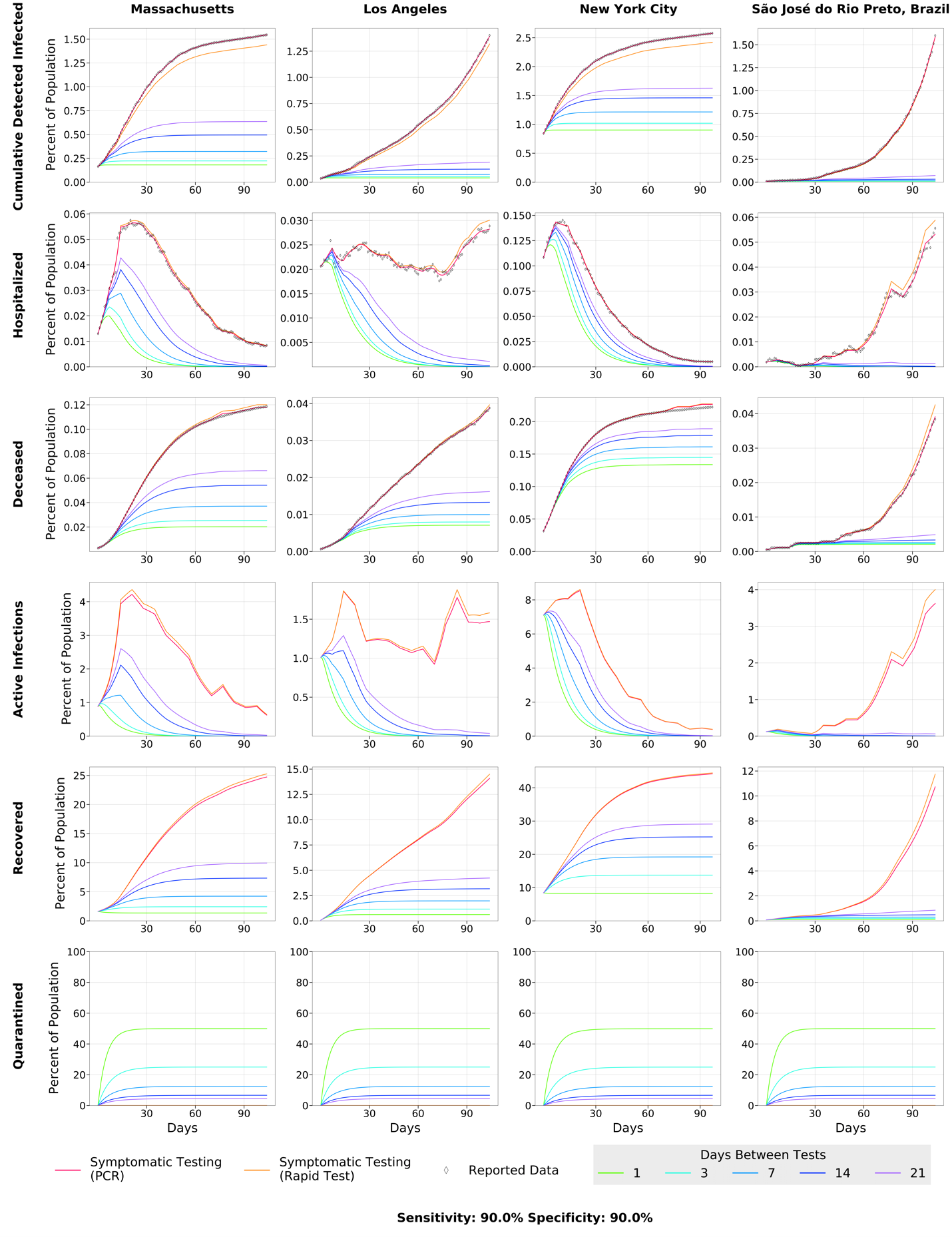

**(B)**
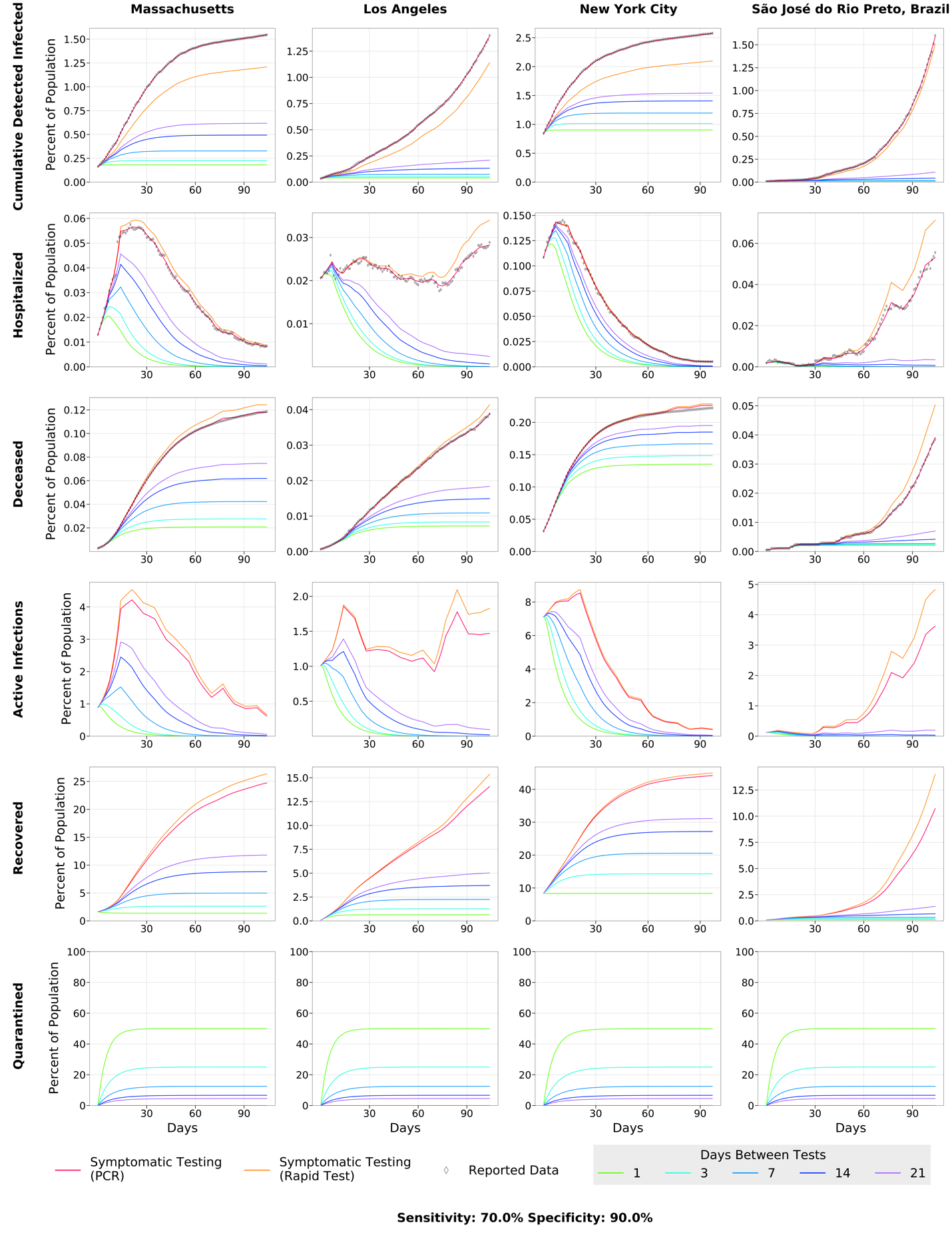

**(C)**

**1**
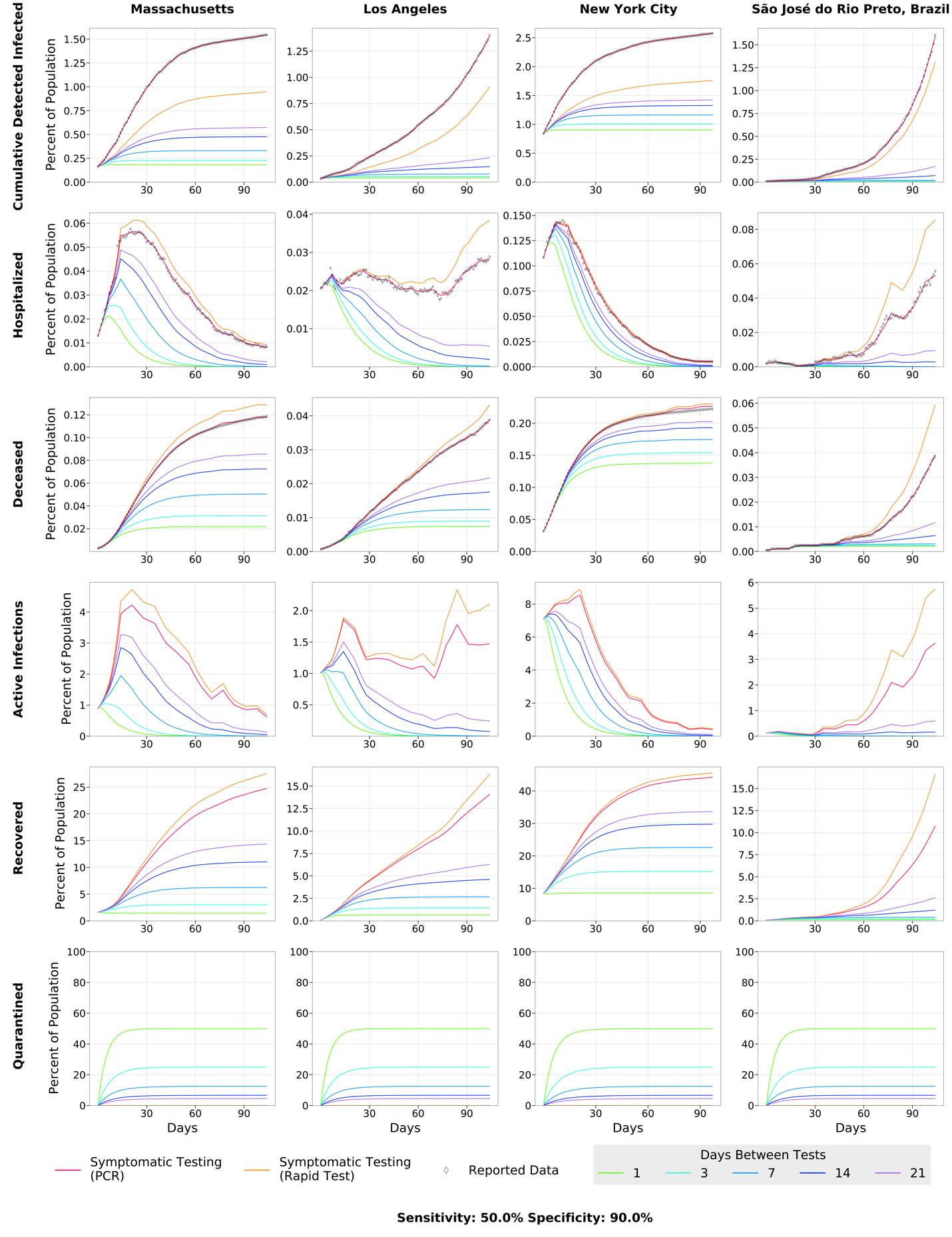

**(D)**
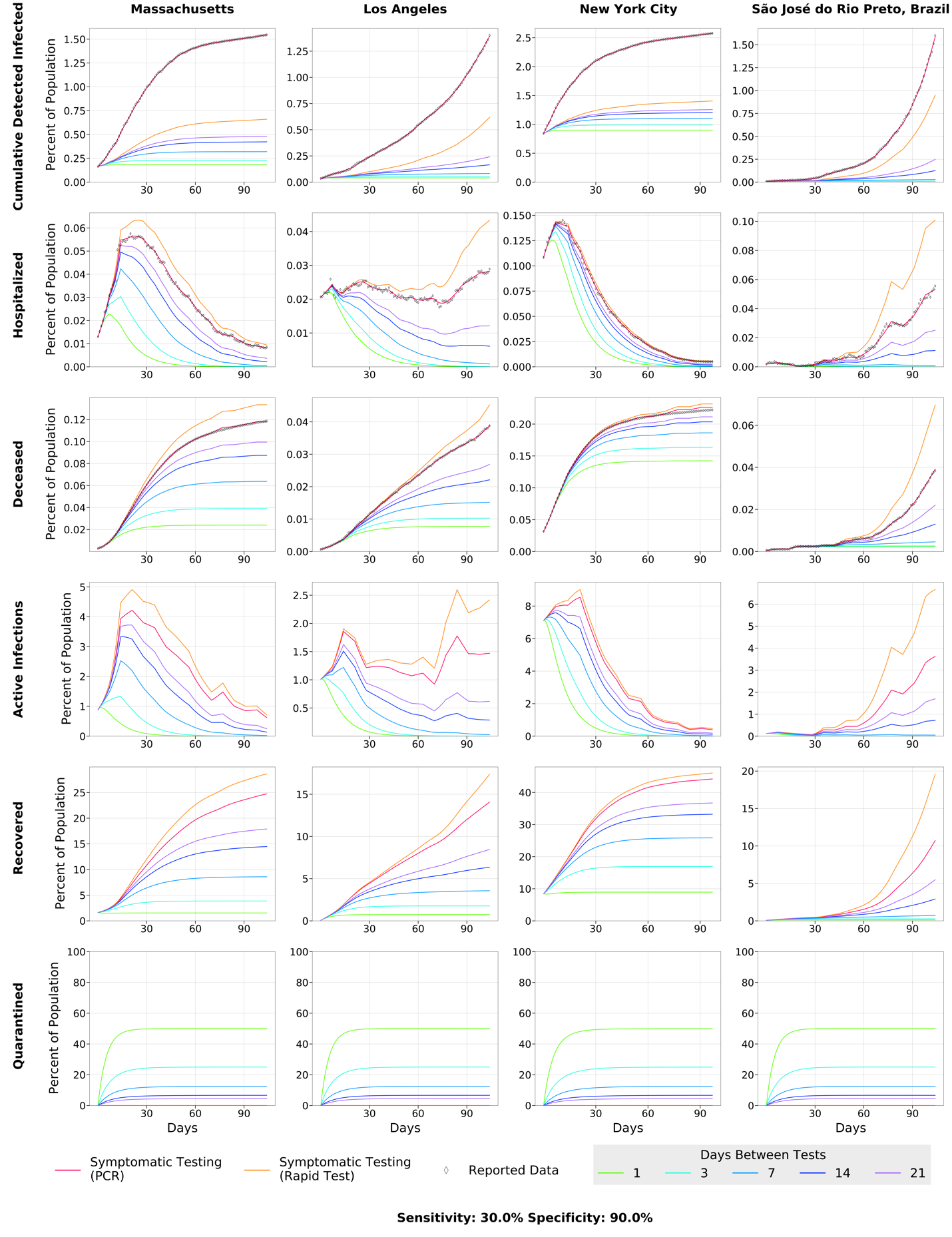

**(E)**

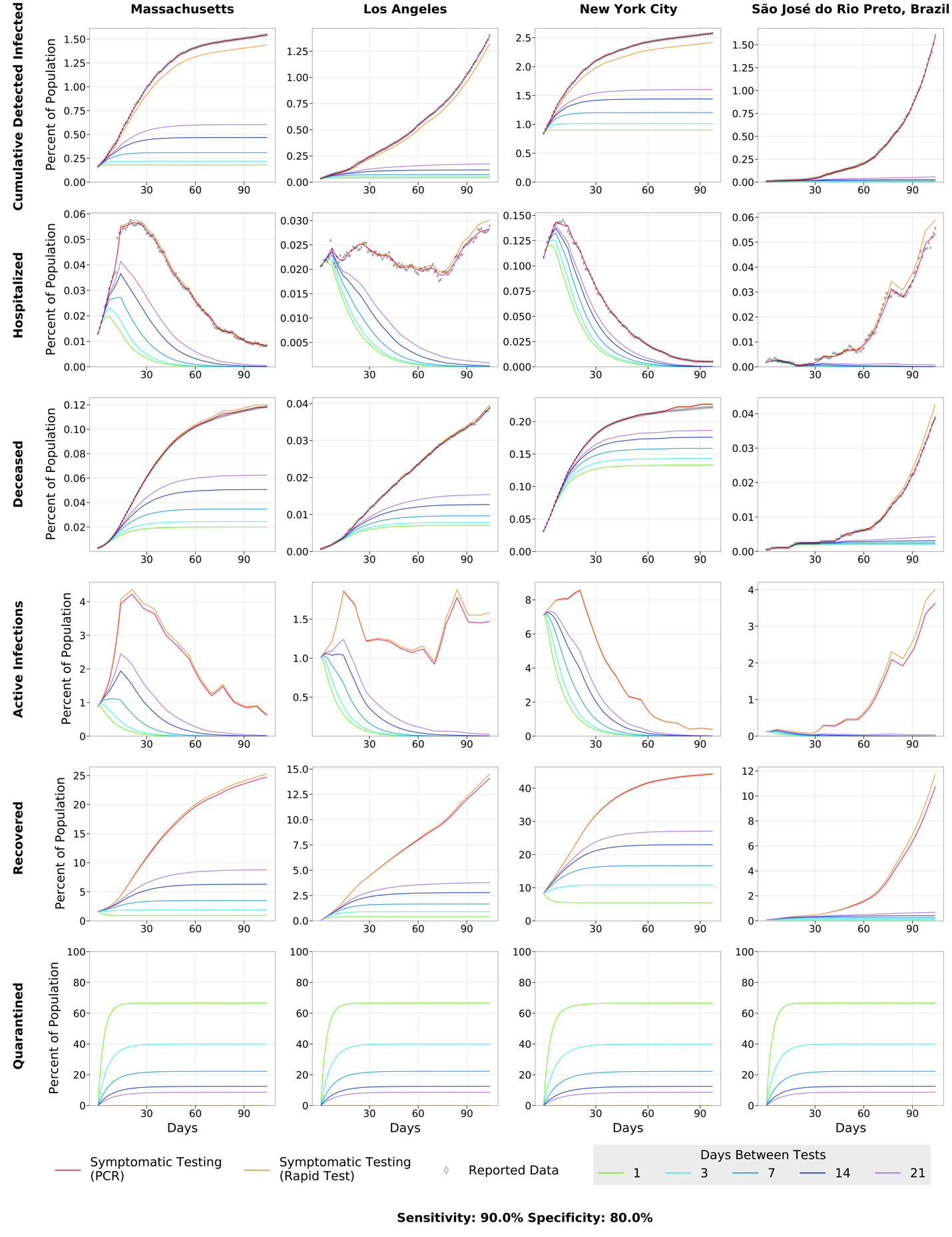

**(F)**

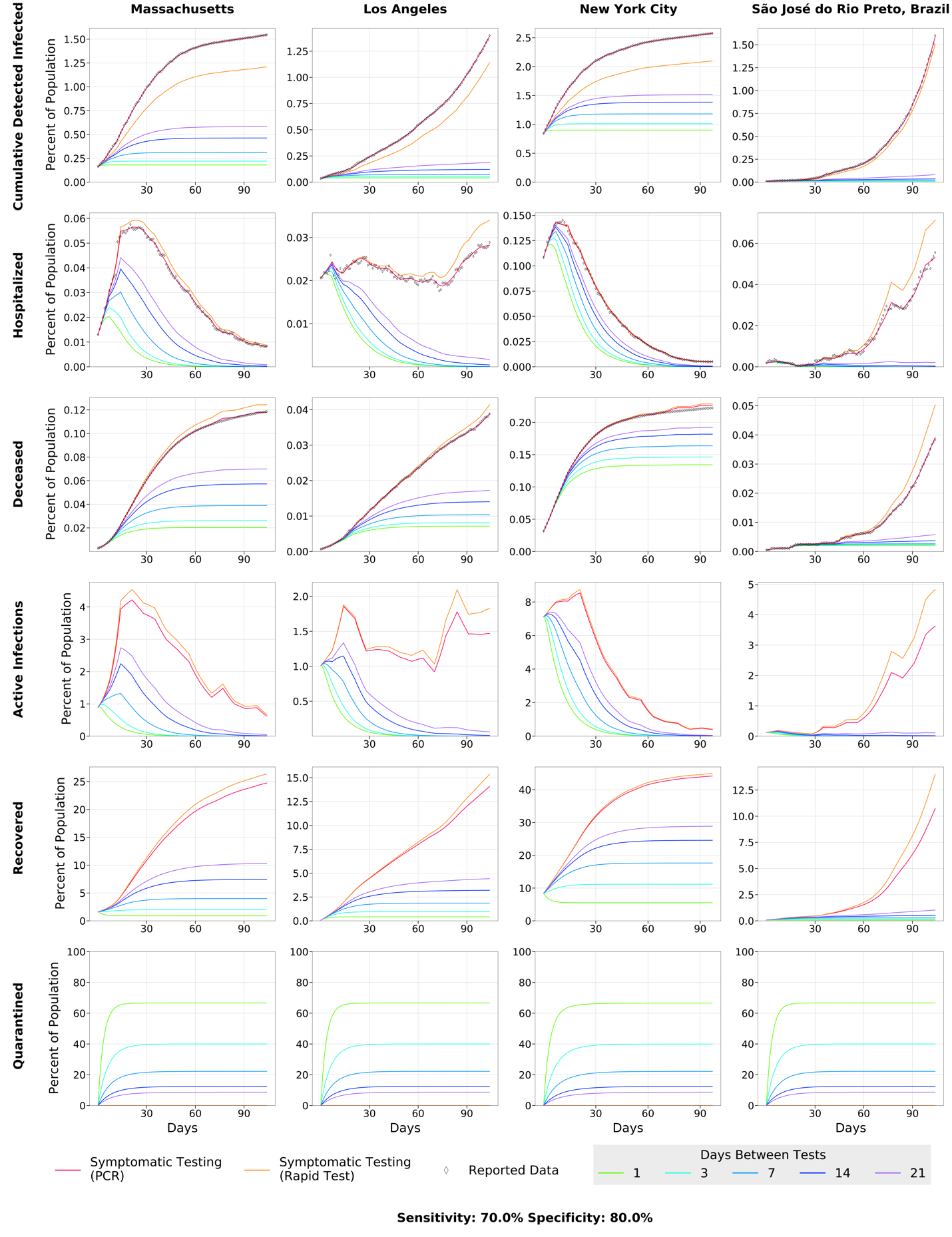

**(G)**

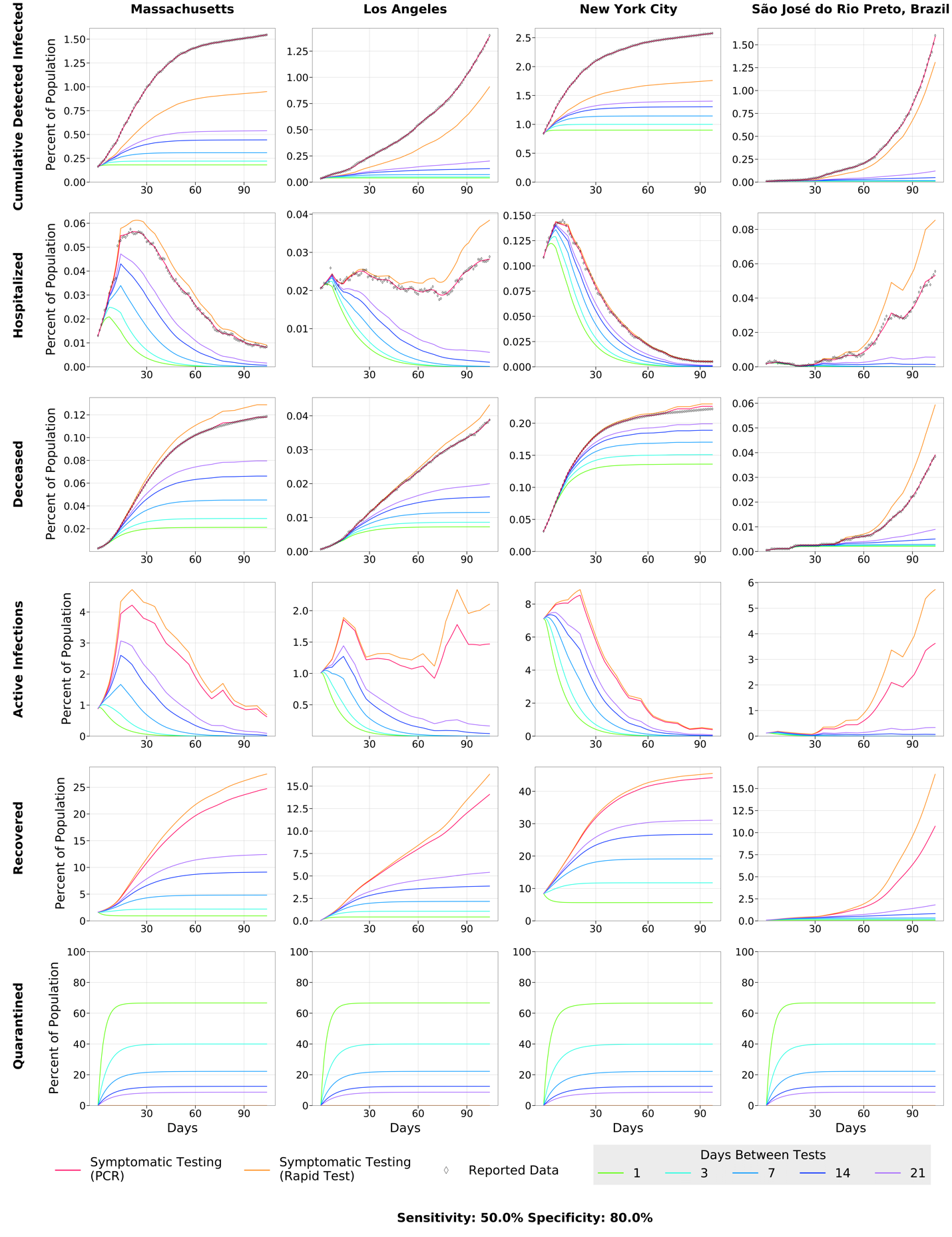

**(H)**

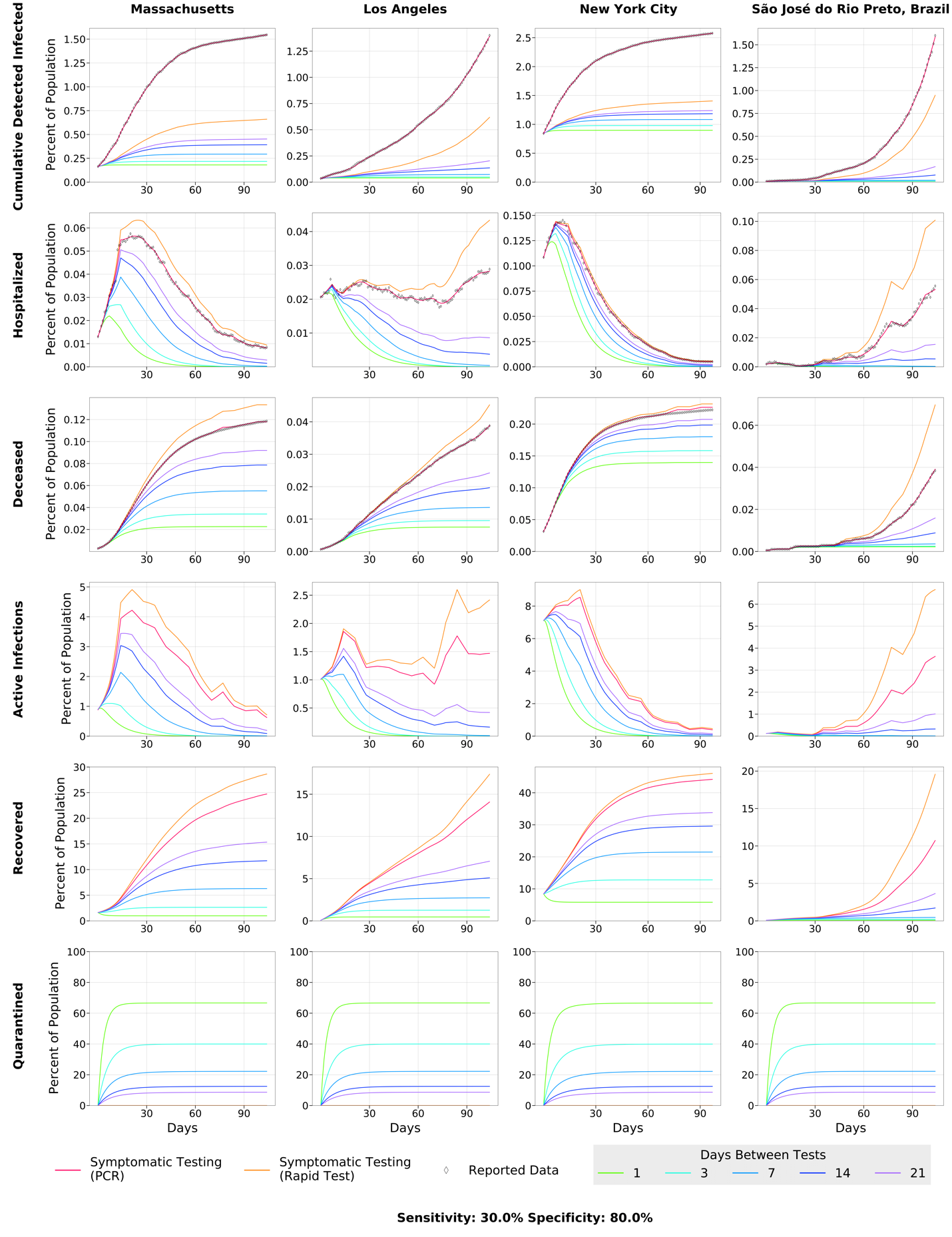

**Supplementary Figure 4. Effect of Rapid Testing Protocol under variable testing sensitivities and increasing frequency under the *SIDHRE-Q* Model.** The Cumulative Infections, Maximum Simultaneously Hospitalized, and Deceased populations are modeled for Massachusetts, Los Angeles, New York City, and São José do Rio Preto in Brazil. The effect of increasing frequency of testing is modeled for various testing sensitivities (30%-90%) with an 80% specificity.

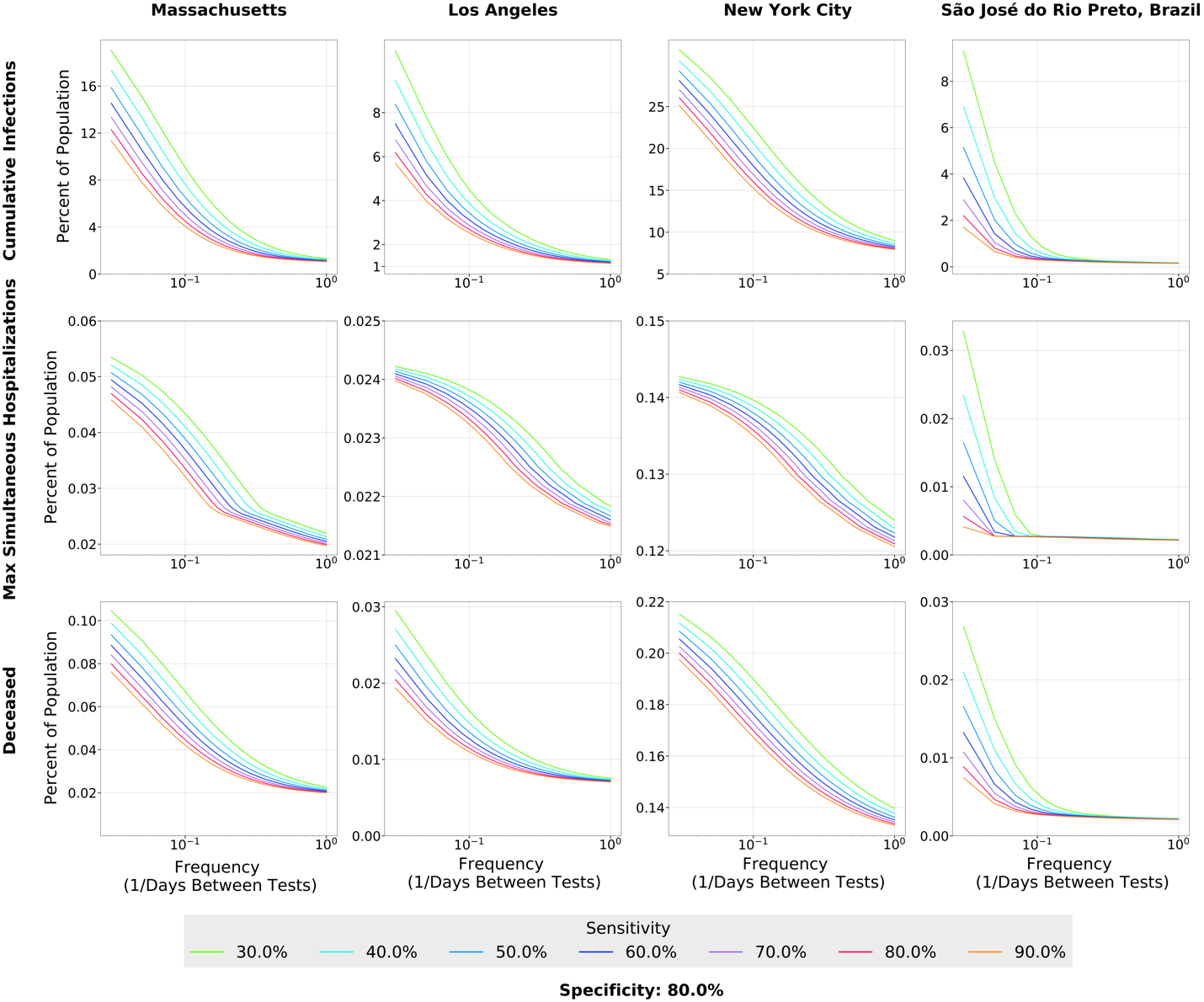

**Supplementary Figure 5.** Missed infections - number of infections that were never diagnosed by the rapid test as a function of log(frequency) for a range of sensitivities with a 90% specificity. Models are shown for MA, LA, NYC, and SJRP.

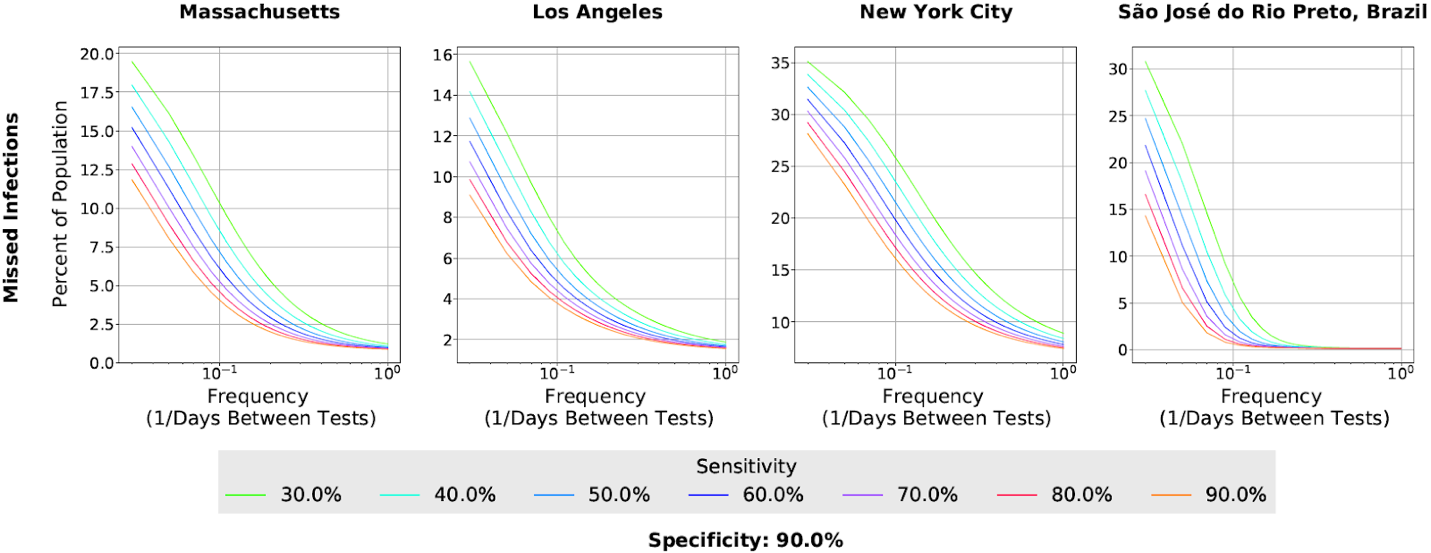

**Supplementary Figure 6**. Time series of the four fitted parameters 𝛼, 𝜈, 𝜇, and 𝜏 (left to right) for MA, LA, NYC, and SJRP (top to bottom). See Table 3 in the Methods section for an explanation of the parameters. The values are extracted every seven days from data provided by the respective regions. The parameters vary significantly over time and location. Flat points occur during the seven day windows where the parameters are held constant. The fitting procedure is also outlined in the Methods section.

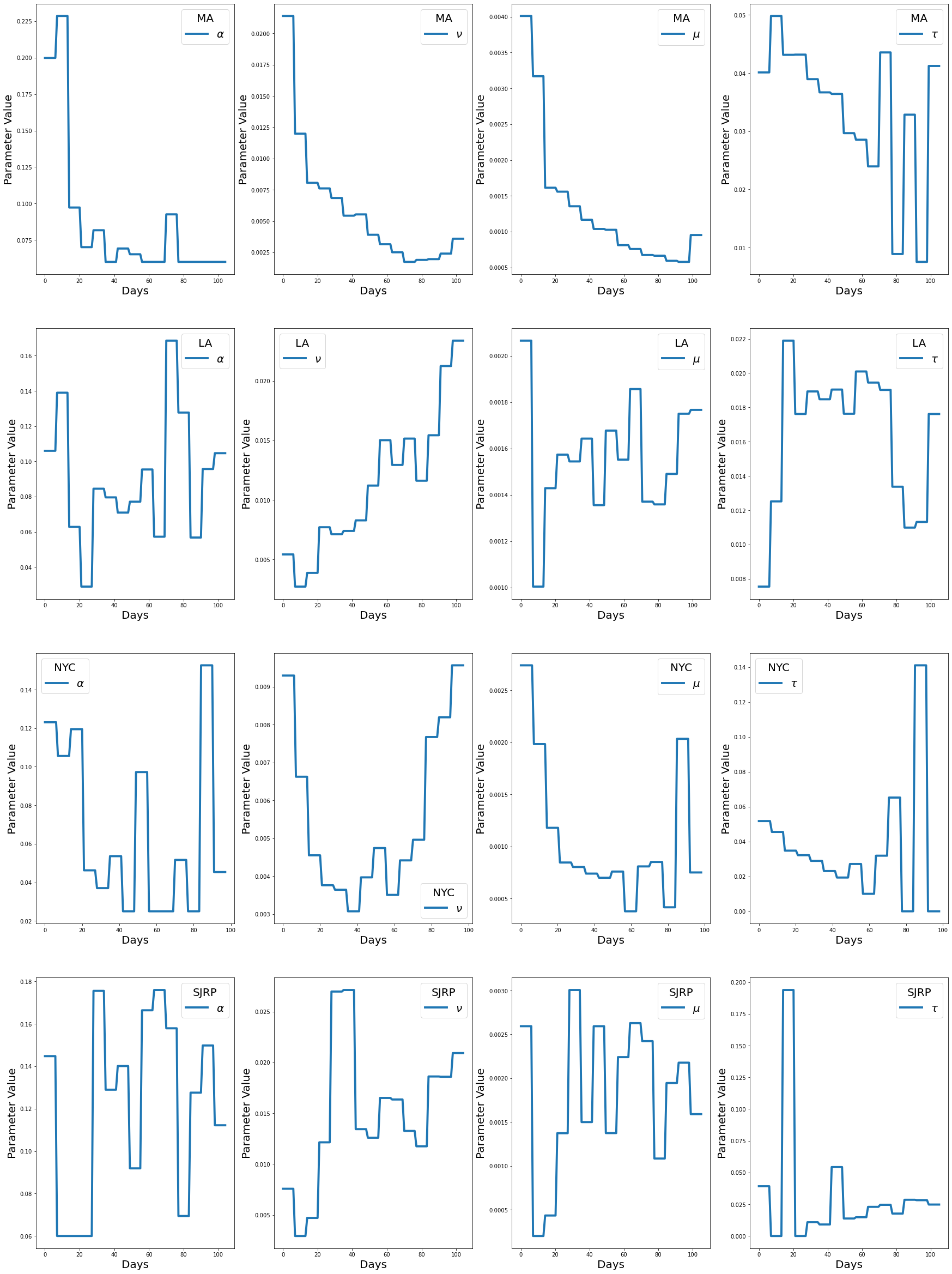

**Supplementary Figure 7.** Time series of the three fitted pieces of data Cumulative Cases, Daily Hospitalized, and Cumulative Deaths (left to right) for each county receiving testing in CA; Ventura (A), Stanislaus (B), Santa Clara (C), San Joaquin (D), San Francisco (E), San Diego (F), San Bernardino (2G), Sacramento (H), Orange (I), Los Angeles (J), Kern (K), Fresno (L), Alameda (M). The counties included satisfy two requirements: population greater than 1.5% of the total CA population and nonzero total number of deaths at each point in time. The fitting procedure is outlined in the Methods section.

**(A)**

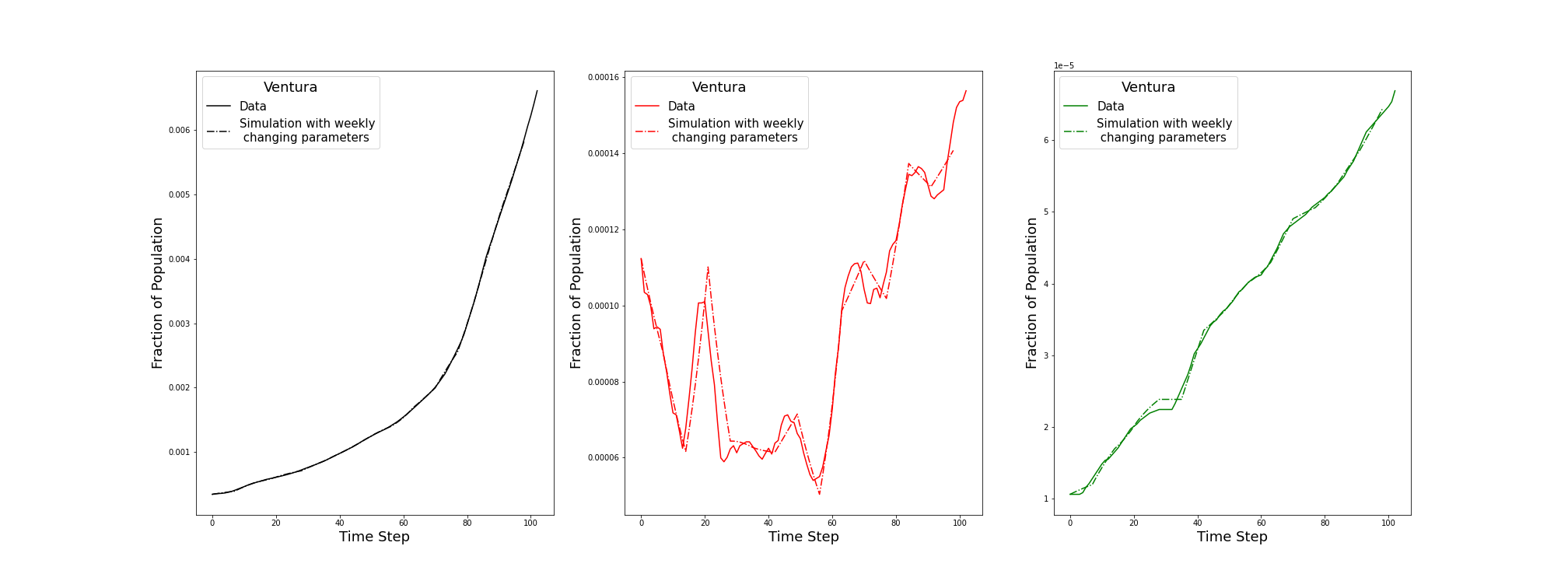

**(B)**

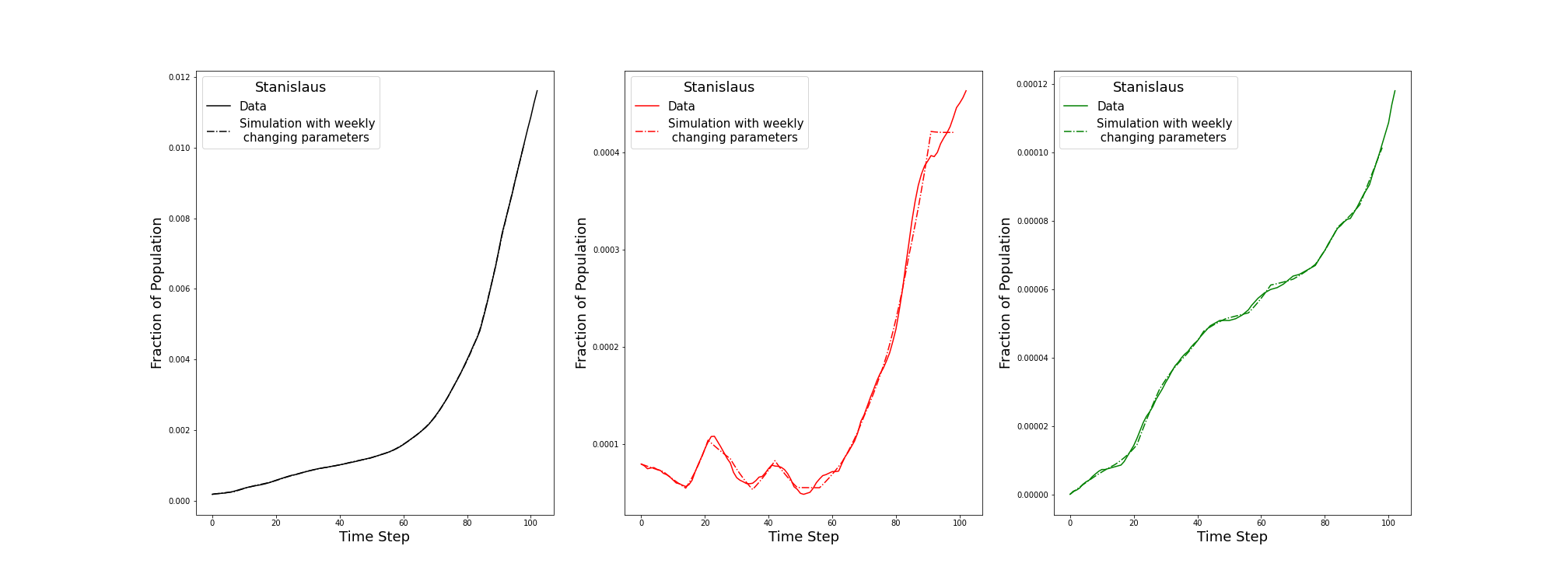

**(C)**

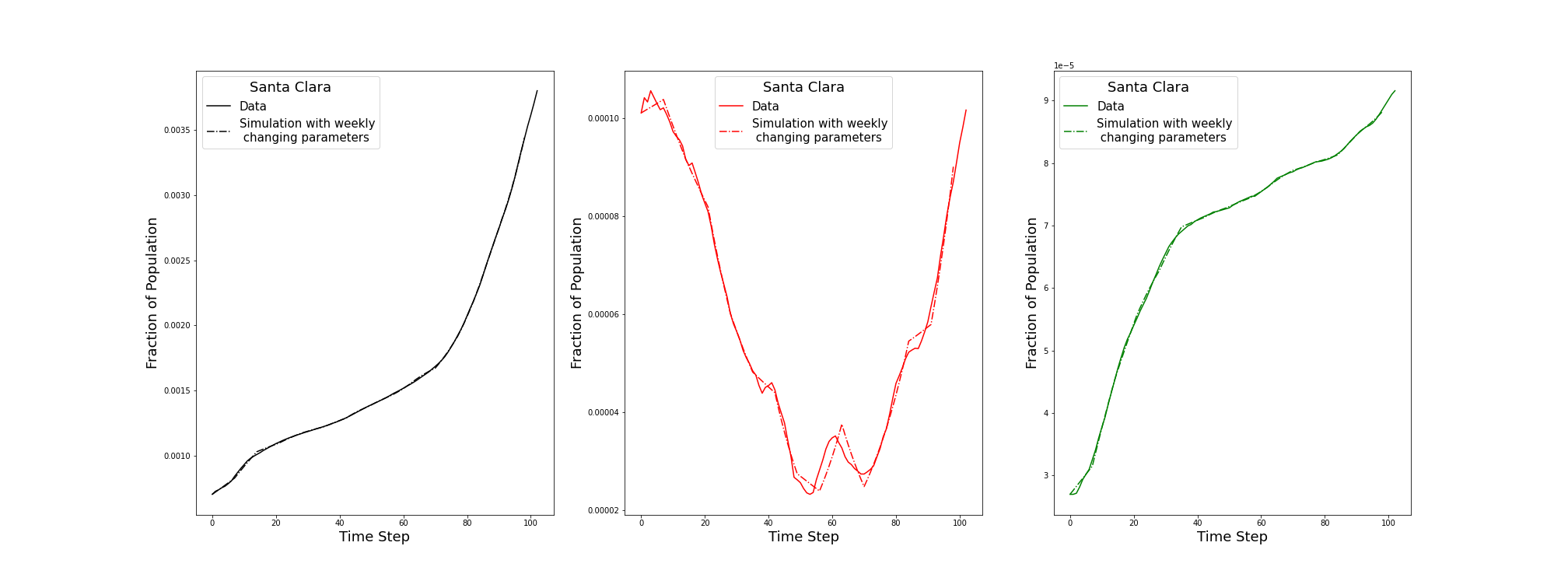

**(D)**

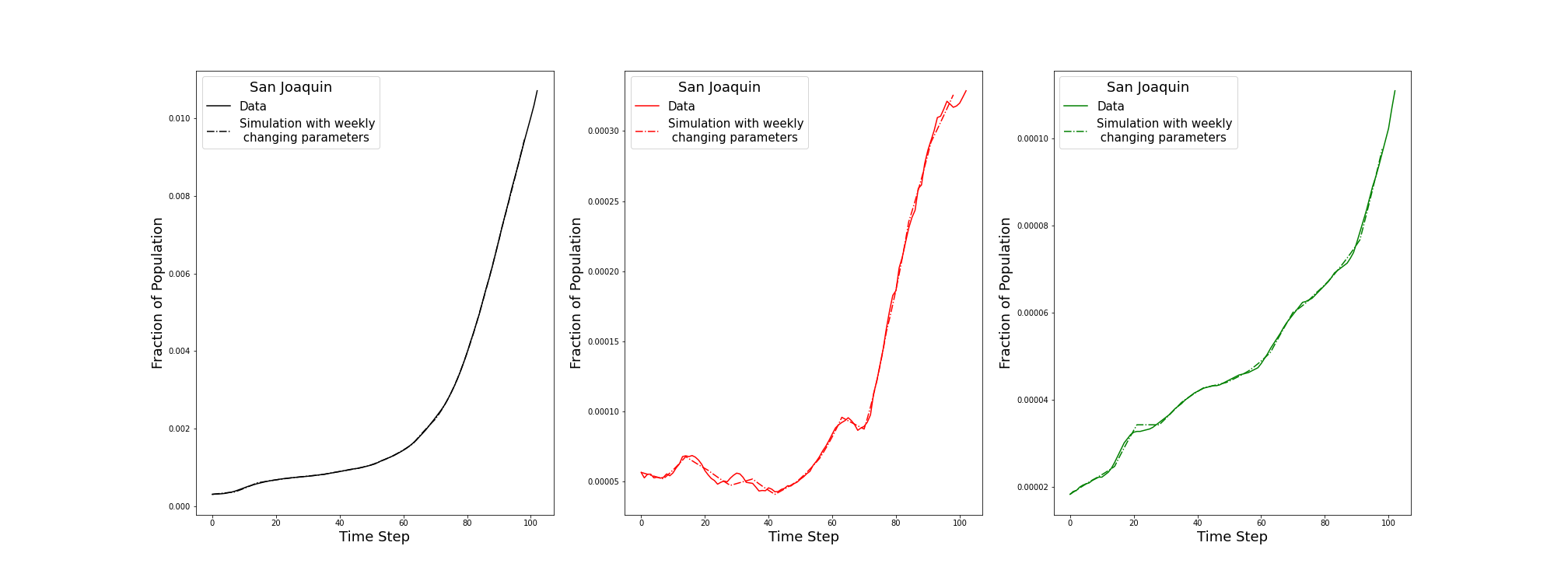

**(E)**

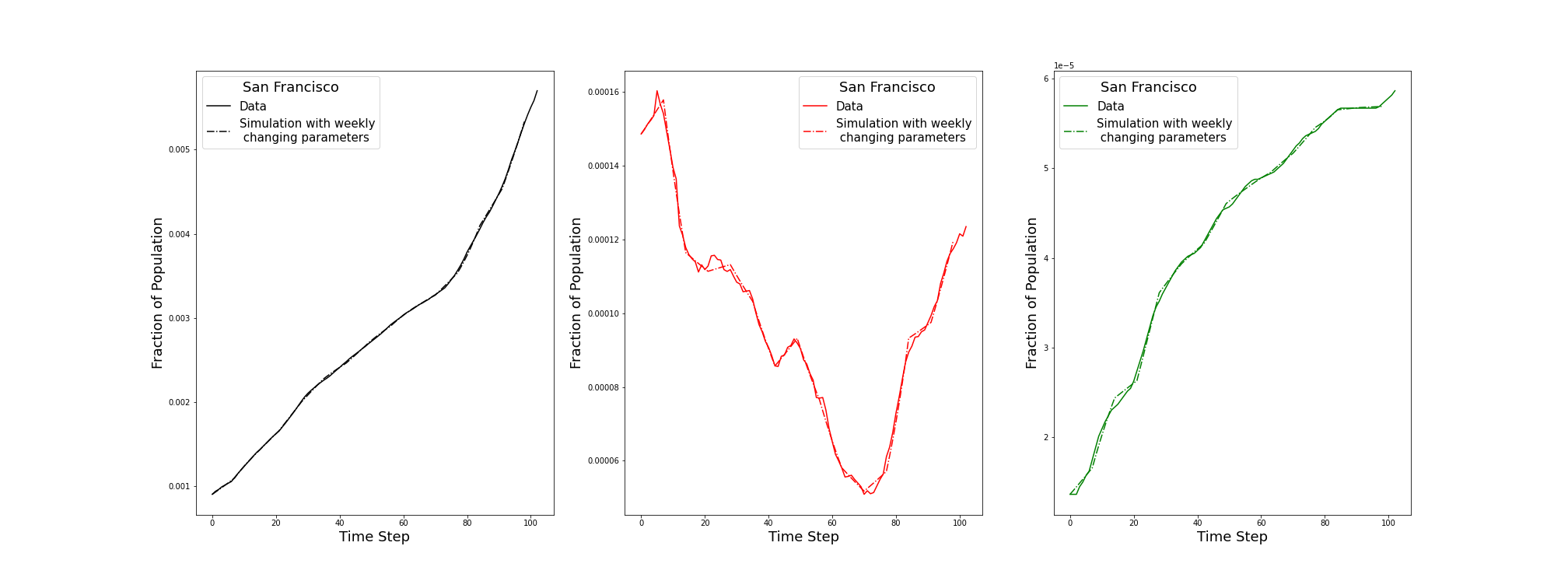

**(F)**

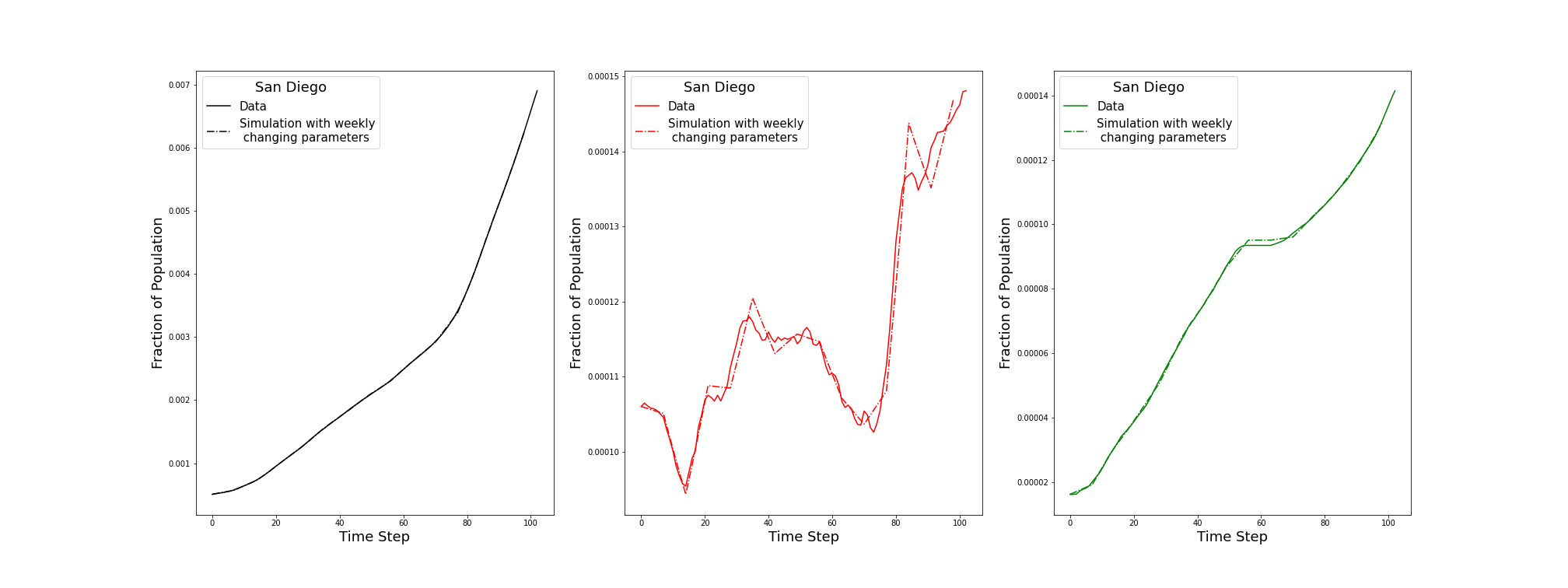

**(G)**

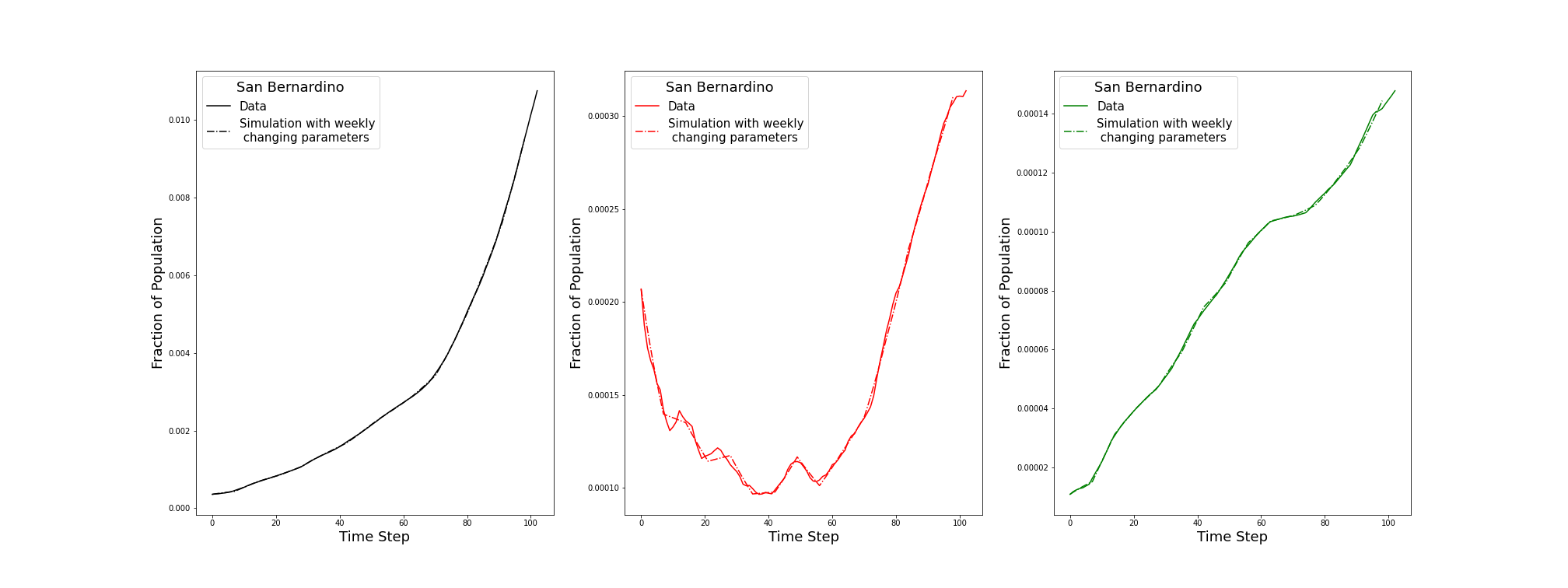

**(H)**

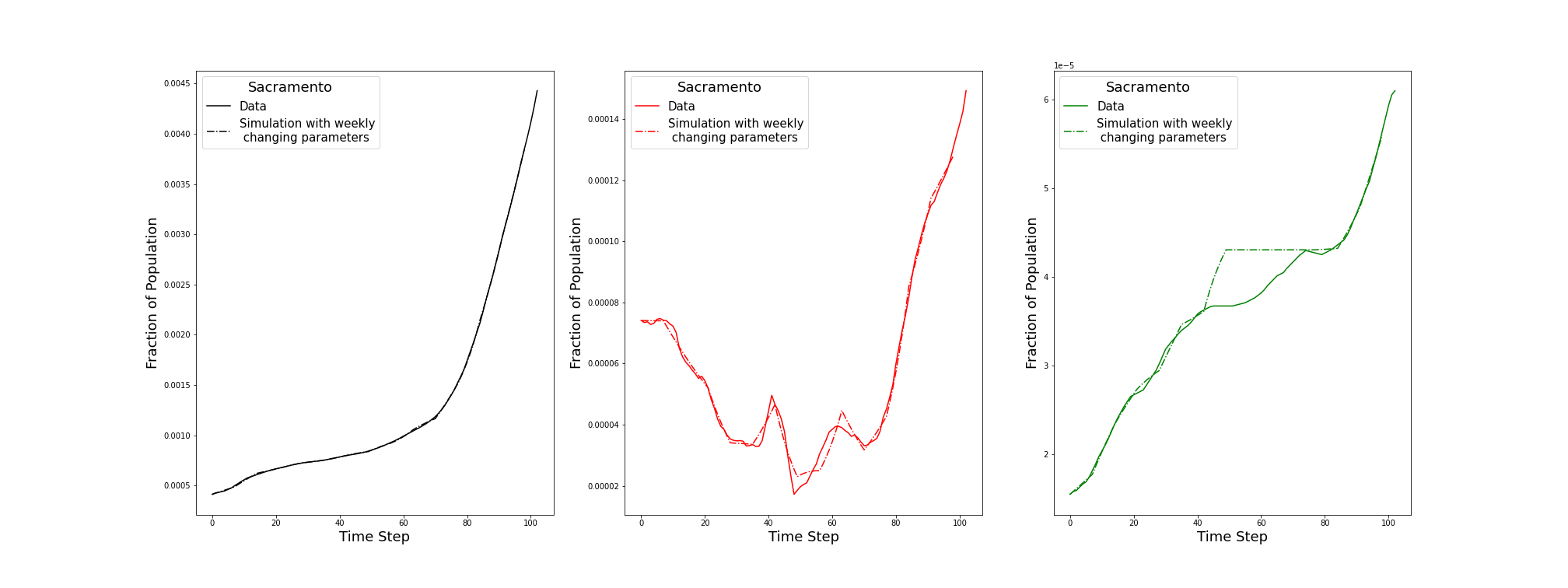

**(I)**

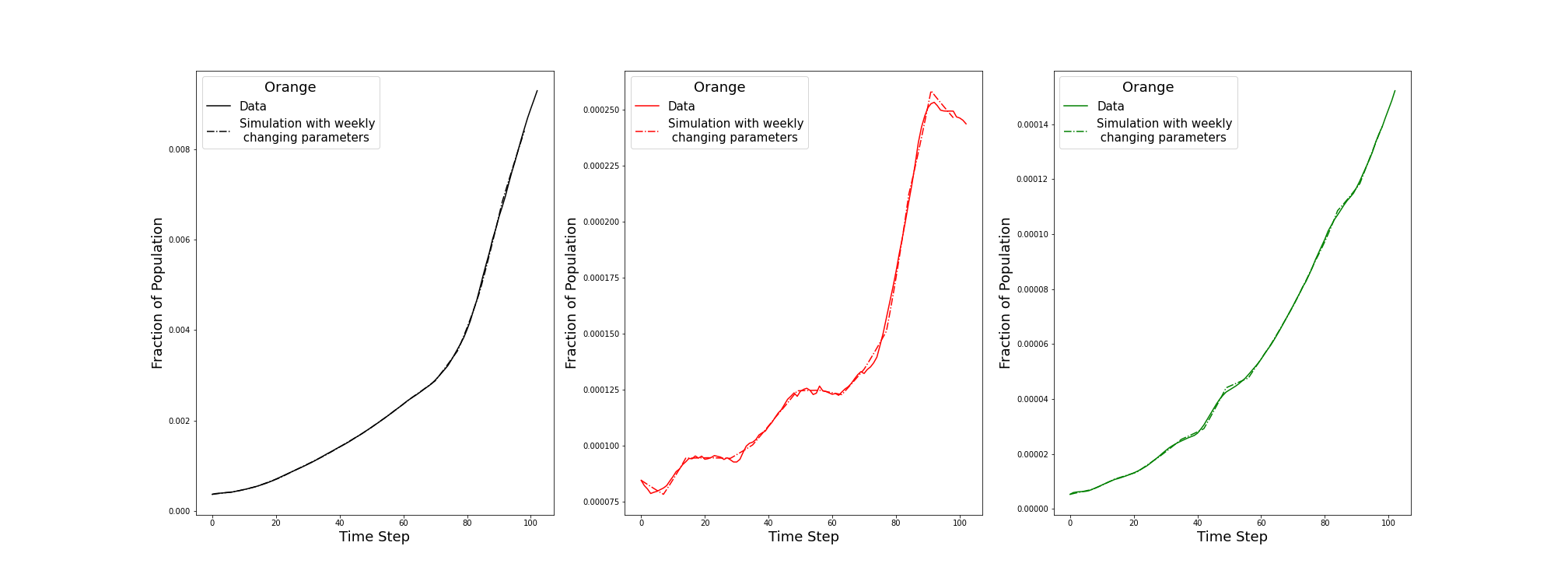

**(J)**

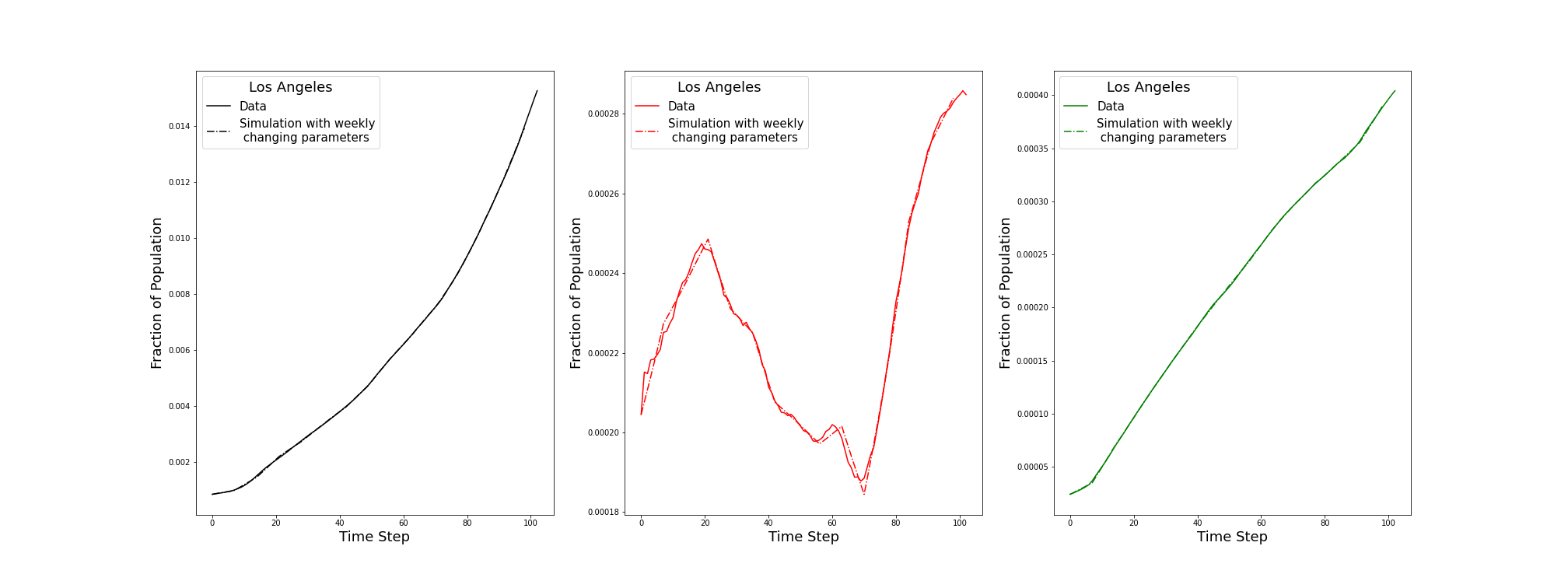

**(K)**

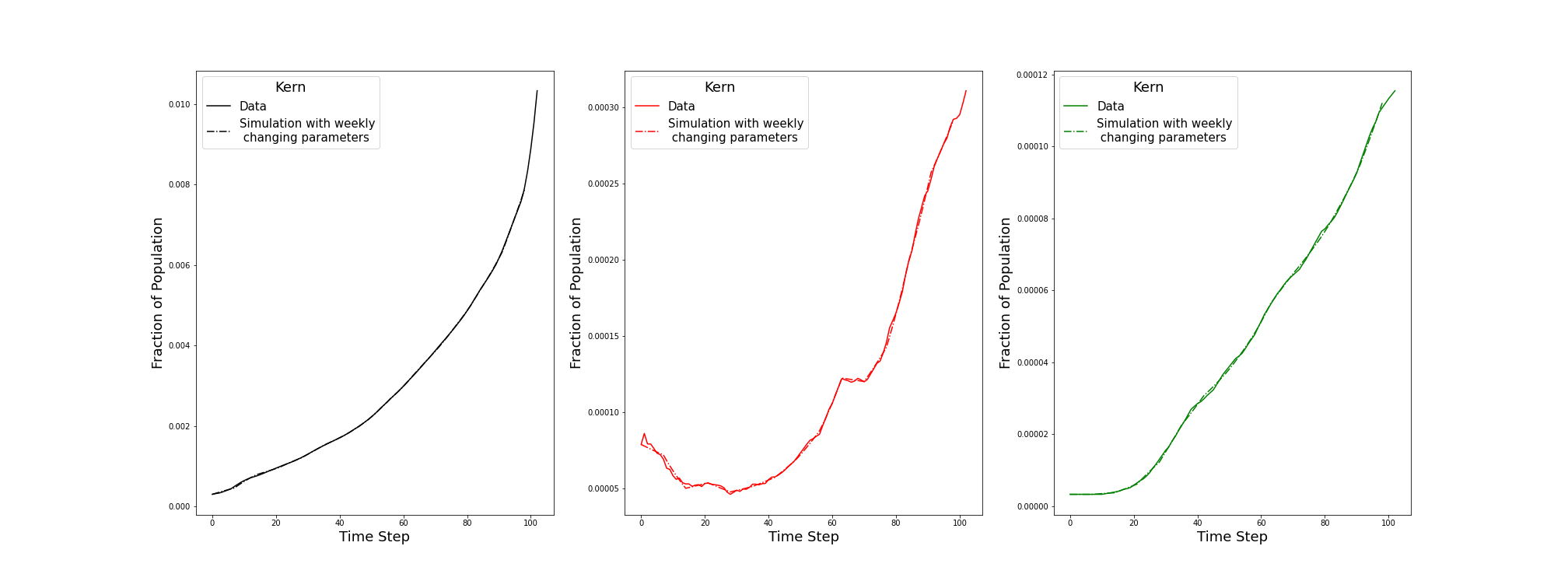

**(L)**

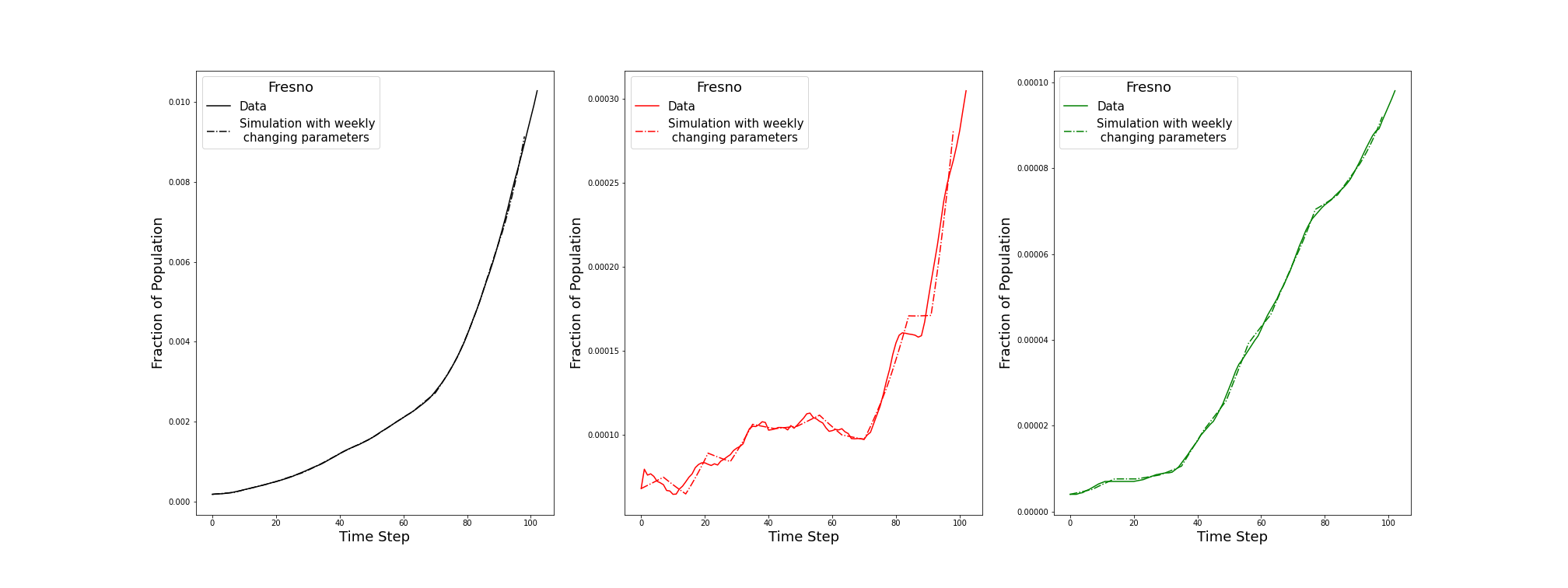

**(M)**

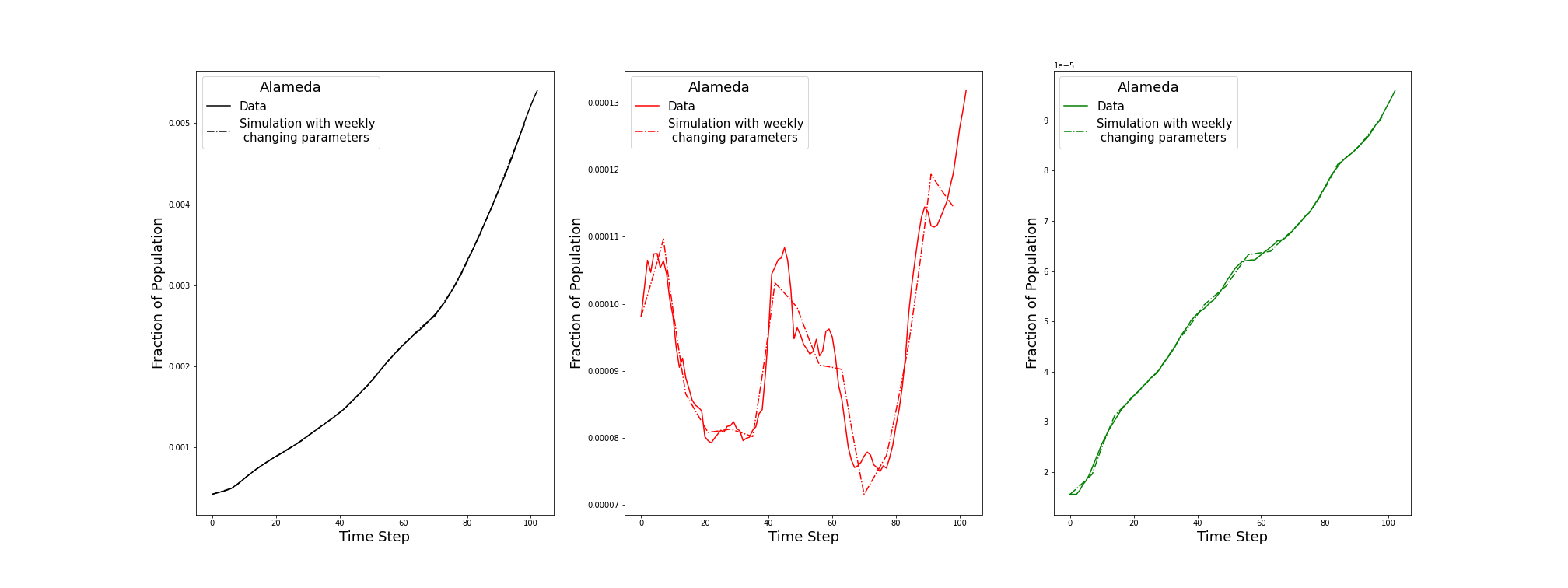

**Supplementary Figure 8.** Dependence of total infections and deaths over the 105 day period in Massachusetts shown as a function of  $\eta$, the value of which indicates quarantine effectiveness, with $\eta$ =0 reflecting full compliance (no transmission due to quarantined individuals) and $\eta$ =1 reflecting no compliance (same transmission due to quarantined individuals as those not quarantined).

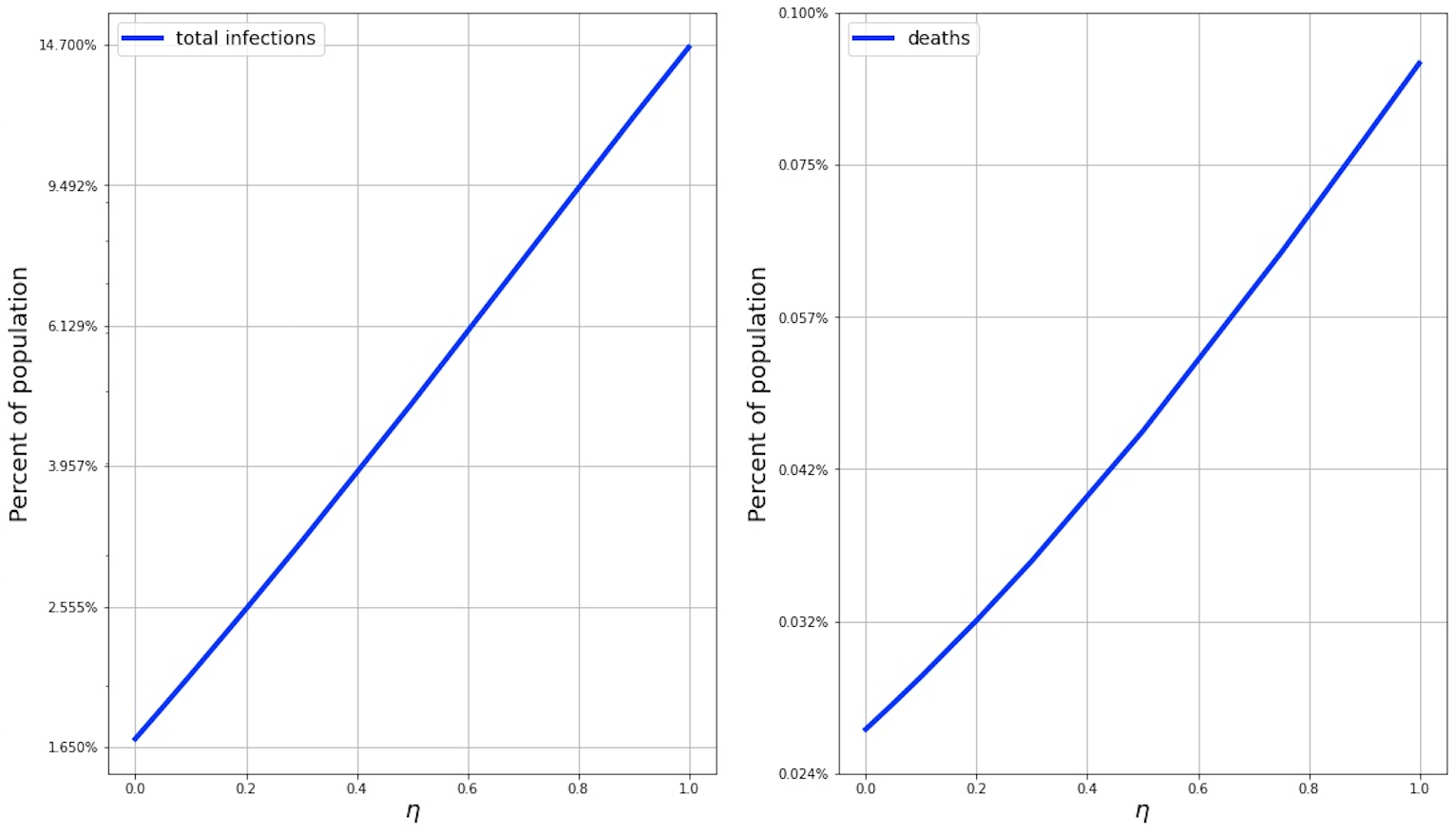

**Supplementary Fig. 9.** Comparison of using a mean value approximation as opposed to fixed duration quarantine and infection periods.  To test the validity of the mean value approximation, we repeat the simulations of the model but replace the single **I** state with 10 substates, each of which corresponds to a different day of infectivity, **D** with 10*10 sub-states, one for each (day infected, day diagnosed) combination, as well as the **Q** state with 10 sub-states corresponding to each day of quarantine.  From sub-state *n < 10* of **I**, there is a flow of value 1 to sub-state *n+1* of **I** as well as a flow into sub-state *n* of **D,** corresponding to rate of diagnosis.  The fixed duration model introduces produces only minimally different results from the mean value scheme when simulated using otherwise identical models.

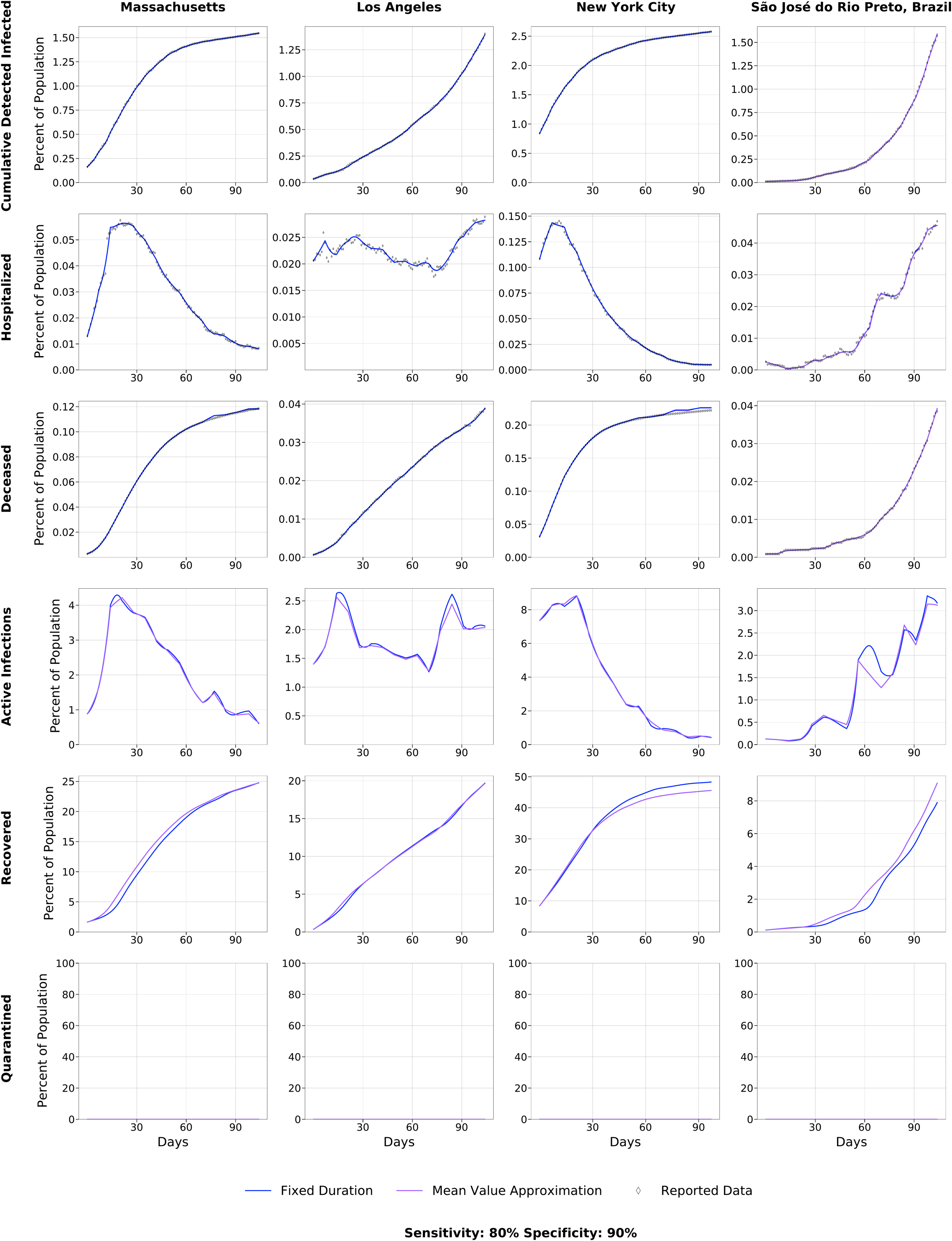
